# Proteomic analysis shows decreased Type I fibers and ectopic fat accumulation in skeletal muscle from women with PCOS

**DOI:** 10.1101/2023.03.08.23286896

**Authors:** E Stener-Victorin, G Eriksson, M Mohan Shrestha, V Rodriguez Paris, H Lu, J Banks, M Samad, C Perian, B Jude, V Engman, R Boi, E Nilsson, C Ling, J Nyström, I Wernstedt Asterholm, N Turner, J T Lanner, A Benrick

## Abstract

**Background:** Polycystic ovary syndrome’s (PCOS) main feature is hyperandrogenism, which is linked to a higher risk of metabolic disorders in women. Gene expression analyses in adipose tissue and skeletal muscle reveal dysregulated metabolic pathways in women with PCOS, but these differences do not necessarily lead to changes in protein levels and biological function.

**Methods:** To advance our understanding of the molecular alterations in PCOS, we performed global proteomic and phosphorylation site analysis using tandem mass spectrometry. Adipose tissue and skeletal muscle were collected at baseline from 10 women with and without PCOS, and in women with PCOS after 5 weeks of treatment with electrical stimulation.

**Results:** Perilipin-1, a protein that typically coats the surface of lipid droplets in adipocytes, was increased whereas proteins involved in muscle contraction and type I muscle fiber function were downregulated in PCOS muscle. Proteins in the thick and thin filaments had many altered phosphorylation sites, indicating differences in protein activity and function. The upregulated proteins in muscle post treatment were enriched in pathways involved in extracellular matrix organization and wound healing, which may reflect a protective adaptation to repeated contractions and tissue damage due to needling. A similar, albeit less pronounced, upregulation in extracellular matrix organization pathways was also seen in adipose tissue.

**Conclusions:** Our results suggest that hyperandrogenic women with PCOS have higher levels of extramyocellular lipids and fewer oxidative insulin-sensitive type I muscle fibers. These could be key factors leading insulin resistance in PCOS muscle while electric stimulation-induced tissue remodeling may be protective.

**Funding:** A.B. holds funding from the Swedish Research Council (2020-02485), E.SV. holds funding from the Swedish Research Council (2022-00550), the Novo Nordisk Foundation (NNF22OC0072904), and I.W.A. holds funding from the Swedish Research Council (2020-01463), Mary von Sydow Foundation, Diabetes Wellness Sverige, and EFSD//European Research Programme on ‘New Targets for Diabetes or Obesity-related Metabolic Diseases’ supported by MSD 2022, and J.N. holds funding from IngaBritt and Arne Lundberg Research Foundation.

## Introduction

Polycystic ovary syndrome (PCOS) is a metabolic and endocrine disorder characterized by clinical signs of hyperandrogenism and reproductive dysfunction (Joham *et al*., 2022). Although not part of the diagnosis, insulin resistance and abdominal obesity are common and lead to an increased risk of type 2 diabetes (Kakoly *et al*., 2019). PCOS affects 8-17% of women worldwide. More than 50% of women with PCOS are overweight or obese, and obesity worsens all symptoms, including insulin resistance (Kakoly *et al*., 2019; Barber & Franks, 2021). In those with PCOS, obesity is associated with an altered adipose tissue function, increased visceral fat, adipocyte hypertrophy, and lower adiponectin levels compared with BMI matched controls (Manneras-Holm *et al*., 2011; Villa & Pratley, 2011; Manneras-Holm *et al*., 2014). In recent independent bidirectional Mendelian studies, obesity is even thought to contribute to or cause the development of PCOS (Brower *et al*., 2019; Zhao *et al*., 2020; Liu *et al*., 2022). Adipose tissue dysfunction also leads to consequences in other metabolic tissues. Overweight/obese women with PCOS are at increased risk of developing lipotoxicity due to increased triglyceride and free fatty acid levels and increased fatty acid uptake into non-adipose cells, including skeletal muscle, which is exacerbated by increased intra-abdominal fat with high lipolytic activity (Dumesic *et al*., 2022). The lipotoxic state in skeletal muscle includes increased expression of genes involved in lipid storage and epigenetic changes, as demonstrated in a PCOS-like sheep model (Guo *et al*., 2020). Consequently, excessive uptake and extracellular storage of free fatty acid in skeletal muscle promotes insulin resistance. Moreover, skeletal muscle from women with PCOS exhibit transcriptional, epigenetic and protein changes associated with insulin resistance, accompanied by an inflammatory, oxidative and lipotoxic state (Corbould *et al*., 2005; Skov *et al*., 2007; Nilsson *et al*., 2018; Stepto *et al*., 2019; Manti *et al*., 2020; Stepto *et al*., 2020; Moreno-Asso *et al*., 2021).

Several studies show that defects in mRNA expression of mitochondrial function, fat oxidation and immunometabolic pathways in adipose tissue and skeletal muscle contribute to insulin resistance in women with PCOS (Skov *et al*., 2007; Nilsson *et al*., 2018; Stepto *et al*., 2019; Manti *et al*., 2020; Stepto *et al*., 2020). Interestingly, the mechanism underlying the reduced insulin-stimulated glucose uptake in PCOS skeletal muscle likely differs from insulin resistance in BMI-matched controls (Corbould *et al*., 2005; Nilsson *et al*., 2018). This finding is consistent with the theory that PCOS is a disorder with genetically distinct subtypes of PCOS (Dapas *et al*., 2020). However, mRNA expression does not always translate to alterations in protein levels and changes in biological function. The first proteomics data on visceral fat and skeletal muscle from severely obese women with PCOS used mass spectrometry of selected protein spots on a 2D gel electrophoresis and were published a decade ago (Corton *et al*., 2008; Montes-Nieto *et al*., 2013; Insenser *et al*., 2016). This method is limited as only a few proteins could be identified. Today, the combination of liquid chromatography with tandem mass spectrometry provides us with thousands of proteins. Recent publications map the proteome in serum, follicular fluid, ovary, and endometrium from women with PCOS, and the proteome in serum may provide new biomarkers for PCOS (Li *et al*., 2020; Abdulkhalikova *et al*., 2022; Wang *et al*., 2022). We here use a nontargeted quantitative proteomics approach to advance our understanding of the pathophysiology in adipose tissue and skeletal muscle from women with PCOS.

We have previously shown that electrically stimulated muscle contractions, known as electroacupuncture, improve glucose regulation and decrease androgen levels in overweight/obese women with PCOS (Stener-Victorin *et al*., 2016). When acupuncture needles are stimulated by low-frequency electrical stimulation, they cause muscle contractions similar to those that occur during exercise. Muscle contractions activate specific physiological signaling pathways, and electrical muscle stimulations and exercise act through partially similar signaling pathways to induce glucose uptake in the acute response to muscle contractions (Benrick *et al*., 2020). In addition, long-term intervention with both exercise and electrical stimulation has been shown to improve glucose regulation, promote ovulation, decrease muscle sympathetic nerve activity and reduce circulating androgens in women with PCOS (Stener-Victorin *et al*., 2009; Hutchison *et al*., 2011; Jedel *et al*., 2011; Johansson *et al*., 2013; Stener-Victorin *et al*., 2016; Tiwari *et al*., 2019). The molecular mechanisms mediating the effect on adipose tissue and skeletal muscle in response to long-term electrical stimulation, remain unclear. Therefore, the second aim is to use proteomics to provide mechanistic explanations for the improved glucose homeostasis in response to long-term electrical stimulation treatment.

## Methods

### Ethical approval

The study was conducted at the Sahlgrenska University Hospital and the Sahlgrenska Academy, University of Gothenburg, Gothenburg, Sweden, in accordance with the standards set by the *Declaration of Helsinki*. Procedures have been approved by the Regional Ethical Review Board of the University of Gothenburg (approval number 520-11) and the study was registered at ClinicalTrials.gov (NTC01457209). All women provided oral and written informed consent before participation in the study.

### Participants and study protocol

Women with PCOS were diagnosed according to the Rotterdam criteria (Group., 2004). This cohort has previously been described in detail and included 21 overweight and obese cases and 21 overweight and obese controls matched for age, weight and BMI (Kokosar *et al*., 2016; Stener-Victorin *et al*., 2016). Of these, subcutaneous adipose tissue and skeletal muscle biopsies from 10 women with PCOS and 10 controls were included in the proteomics analysis. Participants reported to the laboratory in the morning after an overnight fast on menstrual cycle day 1–10 or irrespective of cycle day in ameno-/oligomenorrheic women. Anthropometrics were measured and BMI was calculated as previously described (Kokosar *et al*., 2016; Stener-Victorin *et al*., 2016). Fasting blood samples were taken and HOMA-IR [(fasting insulin (mU/ml) x fasting glucose (mM)/22.5)] was calculated. Circulating testosterone was measured by GC-MS/MS (Kokosar *et al*., 2016). Anthropometrics, reproductive and endocrine variables for those included in the proteomics analysis are given in **Table 1**.

**Table 1.** Anthropometric and biochemical analyses in study participants.

| Variable | Controls baseline (n = 10) | PCOS baseline (n = 10) | Controls vs. PCOS (*P*-value) | PCOS treatment (n = 10) | Baseline vs. treatment (*P*-value) |
| --- | --- | --- | --- | --- | --- |
| Age (years) | 31.2 ± 4.8 | 30.1 ± 5.1 | 0.60 |  |  |
| Weight (kg) | 88.2 ± 11.0 | 88.6 ± 15.6 | 0.82 | 89.0 ± 14.9 | 0.33 |
| BMI (kg/m<sup>2</sup>) | 30.5 ± 3.7 | 30.5 ± 4.1 | 0.88 | 30.7 ± 4.0 | 0.32 |
| Ferriman Gallwey score | 2.1 ± 1.7 | 7.7 ± 4.5 | **0.003** | 9.2 ± 6.0 | 0.48 |
| Testosterone (pg/ml) | 258 ± 77 | 476 ± 211 | **0.004** | 347 ± 220 | **0.012** |
| Insulin (mU/ml) | 8.2 ± 3.5 | 11.4 ± 8.5 | 0.47 | 8.5 ± 4.8 | 0.11 |
| Glucose (mmol/L) | 5.2 ± 0.3 | 4.8 ± 0.4 | **0.044** | 4.8 ± 0.3 | 0.67 |
| HOMA-IR | 1.92 ± 0.96 | 2.75 ± 2.00 | 0.41 | 1.89 ± 1.33 | **0.037** |
| Hemoglobin A1c (mmol/mol) | 31.5 ± 2.6 | 31.7 ± 2.7 | 1.00 | 30.5 ± 2.9 | **0.035** |
| Triglycerides (mmol/L) | 0.71 ± 0.18 | 1.11 ± 0.58 | 0.076 | 1.01 ± 0.58 | 0.066 |

Data are presented as mean ± SD. Differences between PCOS and controls were analyzed by Mann-Whitney *U* test. Wilcoxon signed-rank test was used to analyze changes between measurements at baseline and after 5 weeks of treatment.

Needle biopsies from subcutaneous adipose tissue from the umbilical area and skeletal muscle tissue from vastus lateralis were obtained under local anesthesia (Xylocaine, AstraZeneca AB, Södertälje, Sweden), from cases and controls (**Fig. 1A**). The fat biopsies were rinsed with saline before both tissues were snap frozen in liquid nitrogen and stored at −80°C until further analysis. Thereafter, women with PCOS received low-frequency electrical stimulations causing muscle contractions, so called electroacupuncture. Acupuncture needles were placed in somatic segments corresponding to the innervation of the ovaries and pancreas, bilaterally in abdominal muscle, in quadriceps muscles, and in the muscles below the medial side of the knee. Needles were inserted to a depth of 15–40 mm with the aim of reaching the muscles. Needles were connected to an electrical stimulator (CEFAR ACUS 4; Cefar-Compex Scandinavia, Landsbro, Sweden) and stimulated with a low-frequency (2 Hz) electrical signal for 30 minutes. The intensity was adjusted every 10^th^ minute due to receptor adaptation, with the intention to produce local muscle contractions without pain or discomfort. Treatment was given three times per week over 5 weeks, and the number of treatments varied from 11 to 19. Two sets of needle placements were alternated to avoid soreness (**Fig.1B**) (Stener-Victorin *et al*., 2016). Baseline measurements were repeated after five weeks of treatment, within 48 hours after the last treatment, and new fat and muscle biopsies were collected. After all relevant clinical information was obtained, samples were coded and anonymized.

**Fig. 1.**
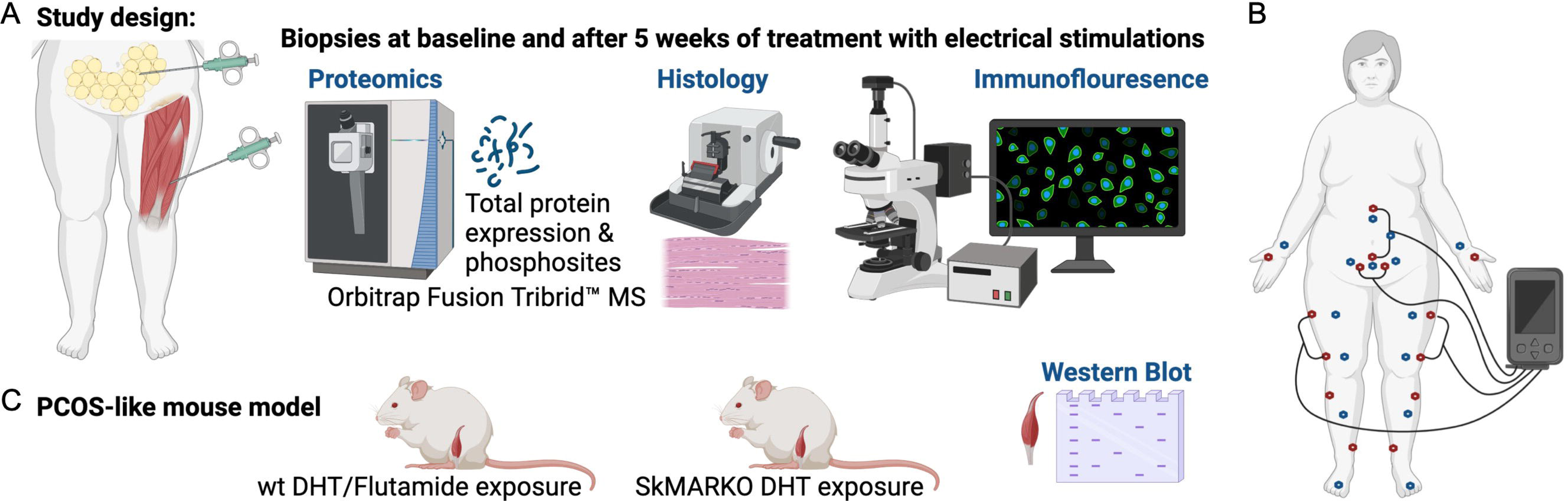
**A)** Muscle and fat biopsies collected from 10 controls and 10 women with PCOS at baseline and after treatment with electrical stimulations. Electrical stimulations were given 3 times/week for 5 weeks. **B)** The electrical stimulation protocol alternating between protocol 1 in red dots and protocol 2 in blue dots. Acupuncture points not connected to the stimulator were stimulated manually. **C)** A PCOS-like mouse model treated with the androgen receptor blocker flutamide or lacking androgen receptors in skeletal muscle (SkMARKO). Created with BioRender.com

### Proteomic sample preparation and LC-MS/MS analysis

#### Tissue Lysis and LC/MS-MS

The individual muscle and adipose tissue biopsies (20-50 mg) were homogenized in 300 μl lysis buffer (50 mM triethylammonium bicarbonate (TEAB; Fluka, Sigma Aldrich) and 2% sodium dodecyl sulfate (SDS)) with 1.4 mm ceramic spheres (FastPrep matrix D) using FastPrep®-24 instrument (MP Biomedicals, OH, USA). The protein concentration was determined using Pierce™ BCA Protein Assay (Thermo Scientific, Rockford, USA) and the Benchmark Plus microplate reader (Bio-Rad Laboratories, Hercules, USA).

#### Sample preparation for global proteomic analysis

Aliquots containing 25 μg of each individual sample were digested with trypsin using the filter-aided sample preparation method. Briefly, protein samples were reduced with 100 mM dithiothreitol at 60°C for 30 min, transferred on 30 kDa MWCO Nanosep centrifugal filters (Pall Life Sciences, Ann Arbor, USA), washed with 8M urea solution and alkylated with 10 mM methyl methanethiosulfonate in 50 mM TEAB and 1% sodium deoxycholate. Digestion was performed in 50 mM TEAB, 1% sodium deoxycholate at 37°C in two stages: the samples were incubated with 250 ng of Pierce MS-grade trypsin (Thermo Fisher Scientific, Rockford, USA) for 3h, then 250 ng more of trypsin was added and the digestion was performed overnight. The peptides were collected by centrifugation, labelled using TMT 10-plex isobaric mass tagging reagents (Thermo Scientific). Sodium deoxycholate was then removed by acidification with 10% trifluoroacetic acid. The mixed labeled samples were fractionated on the AKTA chromatography system (GE Healthcare Life Sciences, Sweden) using the XBridge C18 3.5 μm, 3.0×150 mm column (Waters Corporation, Milford, USA) and 25 min gradient from 7% to 40% solvent B at the flowrate of 0.4 ml/min; solvent A was 10 mM ammonium formate in water at pH 10.00, solvent B was 90% acetonitrile, 10% 10 mM ammonium formate in water at pH 10.00. The initial 40 fraction were combined into 20 pooled fractions in the order 1+21, 2+22, 3+23 etc. The pooled fractions were dried on Speedvac and reconstituted in 20 μl of 3% acetonitrile, 0.1% formic acid for analysis.

#### Sample Preparation for phosphoproteomic analysis

Aliquots containing 450 μg of each individual sample were digested with trypsin using the filter-aided sample preparation method. The phosphopeptides were enriched using Pierce TiO2 Phosphopeptide Enrichment and Clean Up Kit (Thermo Fisher Scientific). The purified phosphopeptide samples were evaporated to dryness, reconstituted in 50 mM TEAB and labelled using TMT 10-plex isobaric mass tagging reagents (Thermo Fisher Scientific). The TMT-labelled phosphopeptide samples were mixed into corresponding sets and purified using Pierce C-18 Spin Columns (Thermo Fisher Scientific). Purified samples were dried on Speedvac and reconstituted in 15 μl of 3% acetonitrile, 0.1% formic acid for analysis.

#### LC-MS/MS Analysis

All samples were analyzed on Orbitrap Fusion Tribrid (Thermo Fisher Scientific) interfaced with Thermo Easy-nLC 1000 nanoflow liquid chromatography system (Thermo Fisher Scientific). Peptides were trapped on the C18 trap column (100 μm X 3 cm, particle size 3 μm) separated on the C18 analytical column (75 μm X 30 cm) home-packed with 3 μm Reprosil-Pur C18-AQ particles (Dr. Maisch, Germany) using the gradient from 5% to 25% B in 45 min, from 25% to 80% B in 5 min, solvent A was 0.2% formic acid and solvent B was 98% acetonitrile, 0.2% formic acid. Precursor ion mass spectra were recorded at 120,000 resolution. The most intense precursor ions were selected (‘top speed’ setting with a duty cycle of 3s), fragmented using CID at collision energy setting of 30 spectra and the MS2 spectra were recorded in ion trap. Dynamic exclusion was set to 30 s with 10 ppm tolerance. MS3 spectra were recorded at 60,000 resolution with HCD fragmentation at a collision energy of 55 using the synchronous precursor selection of the 5 most abundant MS/MS fragments. The phosphopeptides were trapped on the NanoViper C18 trap column (100 μm X 2 cm, particle size 2 μm, Thermo Scientific) and separated on the home-packed C18 analytical column (75 μm X 30 cm) using the gradient from 7% to 32% B in 100 min, from 32% to 100% B in 5 min, solvent A was 0.2% formic acid and solvent B was 80% acetonitrile, 0.2% formic acid. The mass spectrometry settings were the same as described above for the global proteomic analysis, but HCD fragmentation at collision energy 33 was used in MS2.

#### Proteomic data analysis

Identification was performed using Proteome Discoverer version 2.4 (Thermo Fisher Scientific). The database search was performed using the Mascot search engine v. 2.5.1 (Matrix Science, London, UK) against the Swiss-Prot Homo sapiens database. For phosphopeptide samples, phosphorylation on serine, threonine and tyrosine was added as a variable modification. Quantification was performed in Proteome Discoverer 2.4. TMT reporter ions were identified with 3 mmu mass tolerance in the MS3 HCD spectra for the total proteome experiment and with 20 ppm mass tolerance in the MS2 HCD spectra for the phosphopeptide experiment, and the TMT reporter S/N values for each sample were normalized within Proteome Discoverer 2.4 on the total peptide amount. Only the unique identified peptides were considered for protein quantification. The mass spectrometry proteomics data have been deposited to the ProteomeXchange Consortium via the Proteomics Identifications (PRIDE) (RRID:SCR_003411) (Deutsch *et al*., 2020) partner repository with the dataset identifier PXD025358.

The normalized abundance counts were used for the downstream analysis using the Differential Enrichment analysis of Proteomics data package (DEP)(Zhang *et al*., 2018). The counts were log_2_ transformed and proteins that were quantified in less than 2/3 of the samples were removed. Missing values were imputed using random draws from a Gaussian distribution around the minimal value as the missing value was not random but concentrated to proteins with low intensities. The data was batch effect adjusted using ComBat (Johnson *et al*., 2007) with the LC-MS/MS run assigned as the batch covariate. Differential expression analysis was performed on the dataset with DEP’s test_diff function which uses protein-wise linear models and empirical Bayes statistics using limma (Smyth, 2004). The differential expression analysis calculated the log_2_ fold-changes (FC), *p-values* and *q-values* between the three groups control and PCOS at baseline (W0), and PCOS after 5 weeks of treatment (W5) generating two different result datasets. Proteins and phosphorylation enrichment were determined to be significantly differentially expressed between the groups if the *p-value* < 0.05, and log_2_ FC ≥ 0.5 or log_2_ FC ≤ −0.5.

### mRNA expression and DNA methylation arrays

mRNA was extracted from adipose tissue (n=17) and skeletal muscle (n=8) biopsies collected at steady-state during the hyperinsulinemic-euglycemic clamp before and after 5 weeks of electrical stimulation in those with PCOS using the RNeasy Lipid Tissue Mini Kit for adipose tissue and Rneasy Fibrous Tissue Mini Kit for skeletal muscle (Qiagen). Nucleic acid concentration was measured with a spectrophotometer (NanoDrop, Thermo Scientific), and RNA quality was determined with an automated electrophoresis station (Experion, Bio-Rad). A HumanHT-12 v4 Expression BeadChip array (Illumina) was used to analyze global mRNA expression. cRNA synthesis, including biotin labeling, was carried out using an Illumina TotalPrep RNA Amplification Kit (Life Technologies & Invitrogen). Biotin-cRNA complex was then fragmented and hybridized to the probes on the Illumina BeadChip array before hybridized and stained with streptavidin-Cy3 according to the manufacturer’s instructions. Probes were visualization with an Illumina HiScan fluorescence camera. The Oligo package from Bioconductor was used to compute robust multichip average expression measures (Bolstad *et al*., 2003).

For methylation array studies, DNA was isolated from adipose tissue (n=17) and skeletal muscle (n=9) biopsies taken at steady-state during the hyperinsulinemic euglycemic clamp before and after 5 weeks of electrical stimulation of women with PCOS using the QIAamp DNA Mini Kit (Qiagen). Nucleic acid concentrations and purity were estimated with a NanoDrop spectrophotometer (Thermo Scientific, Wilmington, DE), and DNA integrity was checked by gel electrophoresis. Genome-wide DNA methylation was analyzed with the Infinium HumanMethylation450k BeadChip array (Illumina). The array contains 485,577 cytosine probes covering 21,231 (99%) RefSeq genes (Bibikova *et al*., 2011). A DNA Methylation Kit (D5001-D5002, Zymo Research) was used to convert genomic DNA to bisulfite-modified DNA. Briefly, gDNA (500 ng) of high quality was fragmented and hybridized on the BeadChip, and the intensities of the signals were measured with a HiScanQ scanner (Illumina).

#### Array data analysis

The bioinformatics analyses of DNA methylation array data were performed as described previously (Ronn *et al*., 2015). In brief, Y chromosome probes, rs-probes, and probes with an average detection *p*-value > 0.01 were removed. After quality control and filtering, methylation data were obtained for 298,289 CpG sites in adipose tissue and 298,332 CpG sites in skeletal muscle. Beta-values were converted to M-values (M = log2 (β/(1 – β), which were used for all data analyses. Data were then quantile normalized and batch corrected with COMBAT (Johnson *et al*., 2007). The differentially methylated sites were identified using a paired t-test (limma package, Bioconductor). To improve interpretation, after all the preprocessing steps the data were reconverted to beta-values ranging from 0% (unmethylated) to 100% (completely methylated).

### Pathway enrichment analysis

The tool Uniprot, Universal Protein Resource (RRID:SCR_002380), was used to retrieve protein names. We applied enrichment analysis to all differently expressed proteins and phosphorylation sites using Enrichr (RRID:SCR_001575) and STRING (RRID:SCR_005223). Ontology terms with a *q*-value <0.05 and including at least 3 proteins/phosphosites or an odds ratio >100 were considered as enriched.

### Histological analyses and immunofluorescence

Skeletal muscle and adipose tissue biopsies were fixed in histofix (Histolab, Sweden) for >72h and then stored in 70% ethanol. Tissues were dehydrated and fixated in paraffin blocks. Paraffin-embedded adipose tissue and muscle tissue (7 *μ*m) were cut into sections using a rotary microtome (Leica Microtome) and mounted on Superfrost^TM^ Plus Adhesion microscope glass slides (Epredia^TM^ J1800AMNZ, #10149870, Thermo Fisher Scientific). Picrosirius Red staining (cat#24901-250, Polysciences, Inc.) was used to identify and quantify fibrillar collagen in adipose and muscle tissue. Adipose tissue quantification of picrosirius red staining before and after electrical stimulation treatment was performed using a semi-automatic macro in ImageJ software. This macro allows for calculation of the total area (μm2) and the % of collagen staining from each area adjusting the minimum and maximum thresholds. Three different random pictures per section (4-5 sections/subject) were taken at 10x or 20x magnification using a regular bright field microscope (Olympus BX60 & PlanApo, 20x/0.7, Olympus, Japan). All images were analyzed on ImageJ software v1.47 (National Institutes of Health, Bethesda, MD, USA) using this protocol https://imagej.nih.gov/ij/docs/examples/stained-sections/index.html with the following modification; threshold min 0, max 2. Skeletal muscle quantification of picrosirius red staining was performed using the same protocol described above. % of collagen staining was calculated on 8 – 10 images of different microscopic fields from each muscle sample.

For immunofluorescence, the muscle sections were deparaffinized twice in xylene (#534056, Sigma-Aldrich) for 5 min. Sections were rehydrated stepwise twice in 100% ethanol and once in 95%, 70%, 50% ethanol and in deionized water for 5 min each before a final rinse in PBS (#18912-014, Gibco, pH 7.4) for 5 min. The slides were subjected to heat-induced antigen retrieval by heating in antigen retrieval buffer (10 mM Citric acid monohydrate, 0.05 % (v/v) Tween-20, pH 6.0) until it reaches the boiling point and cooling to room temperature. The tissue sections were incubated in blocking buffer (3% normal donkey serum (NDS) (v/v) in PBS) at room temperature for 1 h followed by overnight incubation at 4 °C with primary antibodies (rabbit anti-perilipin-1, dilution 1:150 (Abcam Cat# ab3526) and mouse anti-myosin (skeletal slow), dilution 1:300 (MYH7 antibody, Sigma-Aldrich Cat# M8421) diluted in incubation buffer (PBS containing 0.3% Triton X-100, 1% BSA, 1% NDS and 0.01% sodium azide, pH 7.2). After rinsing the slides three times for 10 min each in PBS, the sections were incubated with fluorochrome-conjugated secondary antibodies (Donkey anti-Rabbit IgG (H+L) Highly Cross-Absorbed Secondary Antibody, Alexa Fluor^TM^ 555, diluted 1:250 (Thermo Fisher Scientific Cat# A-31572) and Donkey anti-Mouse IgG (H+L) Highly Cross-Absorbed Secondary Antibody, Alexa Fluor^TM^ Plus 448, diluted 1:250 (Thermo Fisher Scientific Cat# A32766) diluted in incubation buffer. The muscle sections were rinsed three times for 10 min each in PBS. The slides were mounted with coverslips (#ECN 631-1574, VWR) using Vectashield antifade mounting medium with DAPI (#H-1200, Vectashield). Images were obtained using a Zeiss LSM 700, AxioObserver microscope with Plan-Apochromat 10x/0.45 M27 objective lens. Argon lasers of 488 nm and 555 nm wavelengths were used to excite Alexa Fluor 488 (green) and Alexa Fluor 555 (red) respectively, and Laser Diode 405 nm to excite DAPI (blue). Quantification of perilipin-1 expression in skeletal muscle cells from control and PCOS groups was performed using ImageJ software (National Institutes of Health, Bethesda, MD, USA). The channels of the images were split and converted into 8-bit. The minimum and maximum thresholds were adjusted and kept constant for all the images. Regions of interest were drawn around the cells and empty space for background intensity measurement. The mean perilipin-1 intensity was measured and corrected by deducting the background. A total of 28 PCOS and 33 control cells were quantified. Skeletal muscle fiber size and type were quantified in muscle biopsies frozen in Tissue-Tek® O.C.T. Compound (Sakura Finetek, Gothenburg, Sweden). Cross sections (10 µm) were cryosectioned using a NX70-Epredia cryostat, moved onto glass slides (Expredia, J1800AMNZ) and stored at −20°C. The sections were subsequently immunohistochemically stained for type I fibers and fiber boundaries. In brief, the sections were dried at room temperature for 60 minutes and then fixed in 4% formaldehyde (Merck, 100496) for 30 minutes, permeabilized with 0.5% PBS-Triton™ X-100 (Sigma-Aldrich, 9036-19-5) for 20 minutes and thereafter incubated with 0.25% PBS-Triton™ X-100 with 10% goat serum for 30 minutes. The sections were then incubated with primary MYH7 antibody (1:25; DSHB, BA-F8) for type I fibers overnight at 4°C and subsequently secondary antibody Alexa Flour™ 568 (1:500; Thermo Fisher, A-11031) for 60 minutes at room temperature, both in 0.1% PBS-Triton™ X-100 with 1% BSA. Finally, the sections were incubated with WGA Oregon Green™ 488 (Invitrogen, W7024) for fiber boundaries for 3 hours, whereafter Fluoromount-G™ mounting medium (Thermo Fisher, 00-4959-52) and coverslips were applied. The slides were visualized using a Zeiss AxioScan.Z1 slide scanner. Fiber cross-sectional area was automatically determined using MyoVision v1.0 and the proportion of type I fibers was manually counted on ImageJ. A total of 579 fibers from seven controls (60-150 fibers per muscle section) and 177 fibers (15-80 fibers per muscle section) from women with PCOS were quantified. Data are graphically depicted with each individual fiber quantified.

### Mouse study protocol and Western Blot analysis

All animal experiments were carried out in compliance with the ARRIVE guidelines. 3-week-old wild-type (wt) female mice on C57Bl/6J background were purchased from Janvier Labs (C57BL/6NRj, Le Genest-Saint-Isle, France). Female skeletal muscle androgen receptor knockout mice (SkMARKO) were generated by crossing ARflox mice with B6;C3-Tg(ACTA1-rtTA,tetO-cre)102Monk/J (HSA-rtTA/TRE-Cre) mice (Xiong *et al*., 2022). To induce Cre recombinase expression, at 3 weeks of age, SkMARKO mice were given a diet containing 200 mg/kg doxycycline (Specialty Feeds SF11-059) for the entire duration of the experiment. Mice were maintained under standard housing conditions; ad libitum access to food and water in a temperature- and humidity-controlled, 12-h light/dark environment. Procedures were approved by the Sydney Local Health District Animal Welfare Committee within National Health and Medical Research Council guidelines for animal experimentation or the Stockholm Ethical Committee for Animal Research (approval number 20485-2020). At 4 weeks of age, wt and SkMARKO female mice received a subcutaneous silastic implant containing 5-10 mg DHT (5α-Androstan-17β-ol-3-one, A8380, Sigma-Aldrich, St. Louis, USA) or empty implant (n=5-8/group). A subset of DHT-exposed wt mice received a slow-release flutamide pellet (n=5, 25 mg flutamide/pellet, 90-day release, Innovative Research of America, Sarasota, FL, USA) (**Fig. 1C**) (Ascani *et al*., 2023). At 15-17 weeks of age, the mice were euthanized and gastrocnemius muscle tissue was dissected and snap-frozen.

15-20 mg of gastrocnemius muscle was homogenized in RIPA buffer along with protease inhibitors. Protein was quantified using Pierce BCA assay (Thermo Fisher Scientific, Cat# 23227). Diluted protein lysates were mixed with loading buffer containing β-mercaptoethanol, and heated at 65°C, before being loaded into polyacrylamide gel (handcast 10% or AnykD PROTEAN TGX Precast Protein Stain-Free Gel (Bio-Rad, CA, USA)) and electro-transferred to PVDF membranes using a PVDF Transfer Pack. The membranes were blocked with blocking solution (5% BSA or 5% skim milk in TBS containing 0.1% Tween 20) for 1 h and incubated overnight with anti-myosin primary antibody, dilution 1:1,000 (MYH7 antibody, Sigma-Aldrich Cat# M8421). After washing and incubation with secondary antibody, dilution 1:10,000 (rabbit anti-mouse IgG HRP, Abcam Cat# 97046), immunoreactive protein bands were visualized through enhanced chemiluminescence using ECL substrate. Bands were visualized with the ChemiDoc XRS system (Bio-Rad, CA, USA) and analyzed with the image analysis program Image Lab© (Bio-Rad, CA, USA). After initial imaging, membranes were stripped in mild stripping buffer, blocked, and re-probed with GAPDH (Abcam, Cat# ab8245) or beta-actin antibody (Santa Cruz, Cat# 47778).

### Statistics

Differences in clinical characteristics between women with PCOS and controls were assessed using the Mann-Whitney U test and data are presented as mean ± SD. Wilcoxon signed-rank test was used to analyze changes between measurements at baseline and after 5 weeks of treatment. Differences in protein expression were calculated on log2 fold-change. Proteins and phosphorylation enrichments were determined to be significantly differentially expressed between cases and controls, and after 5 weeks of treatment in women with PCOS, if the *p-value* < 0.05, and log_2_ FC ≥ 0.5 or log_2_ FC ≤ −0.5. Proteomic data are presented as log2 fold-change. Differences between wt controls and treated mice were assessed using the multiple comparisons one-way ANOVA test. A two-way ANOVA was used to analyze the effect of treatment and mouse genotype, and data are presented as mean ± SEM. No statistical methods were used to predetermine sample size, it was based on previous experience. Animals were allocated to experimental groups arbitrarily without formal randomization.

## RESULTS

### Clinical characteristics

Women with PCOS had more antral follicles < 9 mm (22.7 ± 7.9 vs 9.4 ± 4.1, P=0.001), larger ovary volume (8.0 ± 2.9 vs 5.0 ± 2.7 ml, P=0.028), higher Ferriman-Gallwey score, and higher circulating testosterone than controls (**Table 1**). Six of the 10 women with PCOS met all three PCOS criteria; two had hyperandrogenemia and PCO morphology, one had hyperandrogenism and irregular cycles, and one had irregular cycles and PCO morphology. Five weeks of treatment lowered testosterone, HOMA-IR, and HbA1c levels, tended to decrease triglyceride levels but did not improve Ferriman-Gallwey score (**Table 1**).

### Total protein expression and phosphorylation in skeletal muscle

In total, we identified 3480 proteins in skeletal muscle. 58 unique proteins were found to be differentially expressed in skeletal muscle from women with PCOS versus controls (*p* < 0.05, and log_2_ FC ≥ 0.5 or ≤ −0.5, **Fig. 2A, Supplementary file 1a**). 25 proteins were upregulated and 33 were downregulated in women with PCOS and the expression log2 fold change in expression ranged from −3.06 to 1.21 (**Supplementary file 1a**). We searched for enriched signaling pathways among the differently expressed proteins using STRING analysis. Our network had significantly more interactions than expected (enrichment p-value <1e^-16^). This means that the differently expressed proteins have more interactions with each other than what would be expected from a random set of proteins of the same size. Such an enrichment indicates that the proteins are biologically linked. Upregulated proteins were enriched in lipid metabolic pathways including negative regulation of cholesterol transport, regulation of lipoprotein lipase activity, and negative regulation of metabolic processes (**Supplementary file 1b**). This enrichment was driven by increased expression of apolipoprotein C-I, C-II and C-III (**Fig. 2B**), which are also enriched in the negative regulation of lipoprotein lipase activity (GO:0051005). Aldo-keto reductase family 1 members C1 and C3 (*AKR1C1* and *AKR1C3,* **Fig. 2B**), which have an androsterone dehydrogenase activity (GO:0047023), were also upregulated and AKR1C1 was strongly correlated to higher circulating testosterone levels (Spearman rho=0.65, *P*=0.002), suggesting that muscle may produce testosterone via the backdoor pathway. Moreover, perilipin-1 that typically coats the surface of lipid droplets in adipocytes (Gandolfi *et al*., 2011; Zhao *et al*., 2021), so-called extra-myocellular adipocytes, was increased in PCOS muscle. The increased expression of perilipin-1 was confirmed by immunofluorescence staining and quantification of muscle biopsies (**Fig. 2C, D**).

**Fig. 2.**
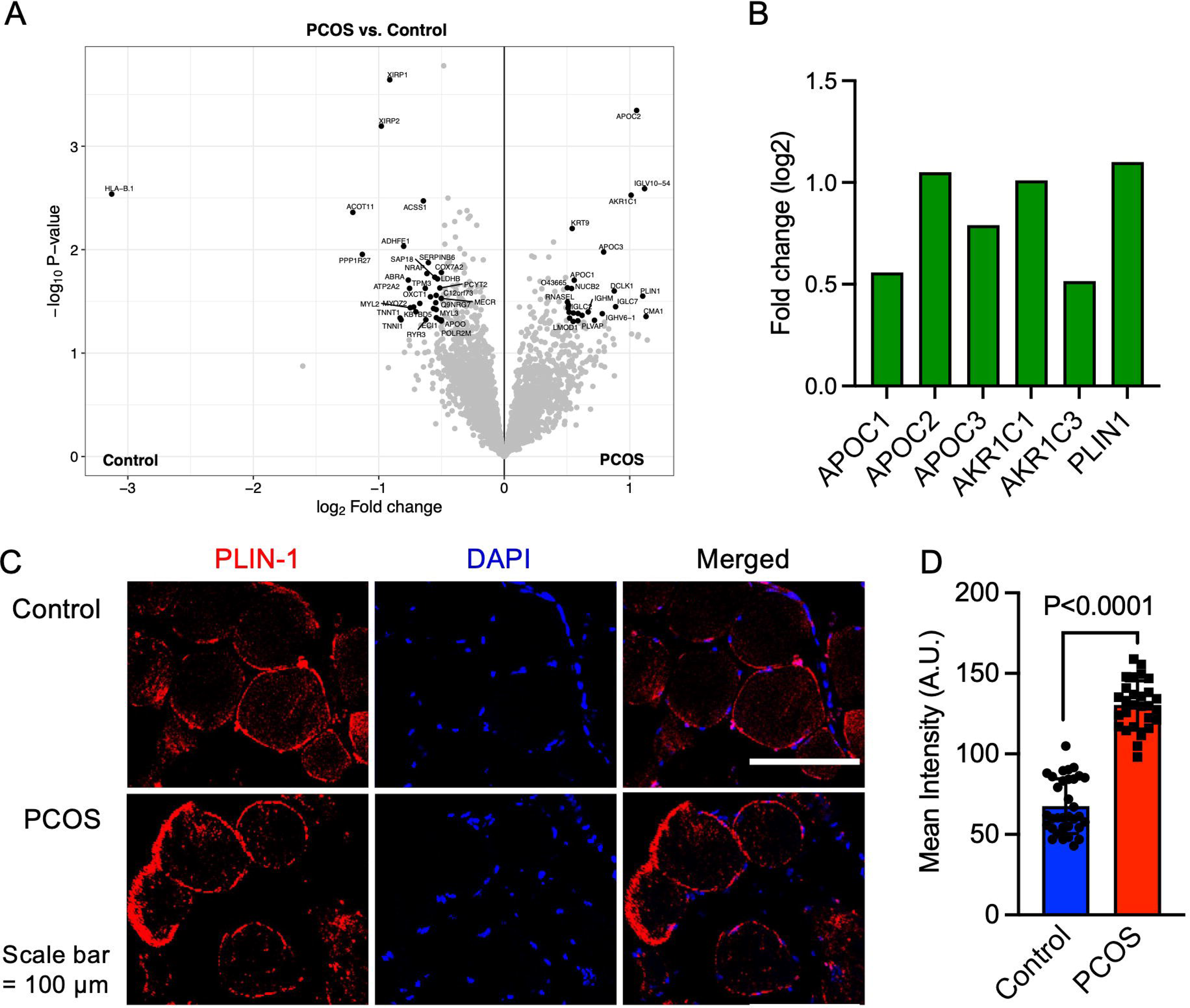
**A)** Volcano plot showing the mean protein log2 fold change in skeletal muscle (PCOS vs controls) plotted against the −log10 *p* value highlighting significantly regulated proteins in black (*p*<0.05, log2 fold change ± 0.5), n=10/group. **B)** Increased protein expression of apolipoproteins C1 and C2, aldo-keto reductase (AKR) family 1 C1 and C3, and perilipin-1 in those with PCOS, **C)** staining of perilipin-1 and DAPI in skeletal muscle, **D)** quantification of perilipin-1 staining in skeletal muscle cells from control and PCOS groups.

The down-regulated proteins in PCOS were enriched in pathways involved in muscle contraction, actin filament organization, and transition between fast and slow fibers **(Fig. 3A)**. All significantly enriched pathways are listed in **Supplementary file 1b.** Expression of myosin heavy chain beta, which is specific for type I muscle fibers was decreased in PCOS (**Fig. 3B**). Several proteins that are more highly expressed in type I muscle fibers, consistently had a lower expression in women with PCOS, for example, myosin heavy chain 7, myosin regulatory light chain 2 and 3, troponin I and troponin T in slow skeletal muscle, and the Ca^2+^ pump SERCA2a. These are proteins located in both the thick filaments (myosin regulatory light chains) and the thin filaments (troponins) of slow-twitch fibers (**Fig. 3C**). A decrease in type I slow-twitch muscle fibers was also supported by staining of human myosin heavy chain 7 (MYH7) as a marker (**Fig. 3D**). To further assess whether there was a reduced fiber size or decreased number of type I fibers; fiber cross-sectional area of the fibers and the percentage of type I fibers were analyzed. The quality of muscle biopsies was impaired in the PCOS group; therefore, 60% fewer fibers from each individual were analyzed in the PCOS group compared with controls (*P*=0.02). There was no significant difference in the mean cross-sectional area of the fibers (4530±720 in controls versus 4281±902 μm in PCOS, *P*=0.64, n=5-7/group) or the percentage of type I fibers (48±12 versus 45±17 μm, *P*=0.69, n=4/group) in the relatively few individuals analyzed (**Fig. 3E**). There was, however, a decrease in the individual fiber cross-sectional area in PCOS muscle versus controls (**Fig. 3F**).

**Fig 3.**
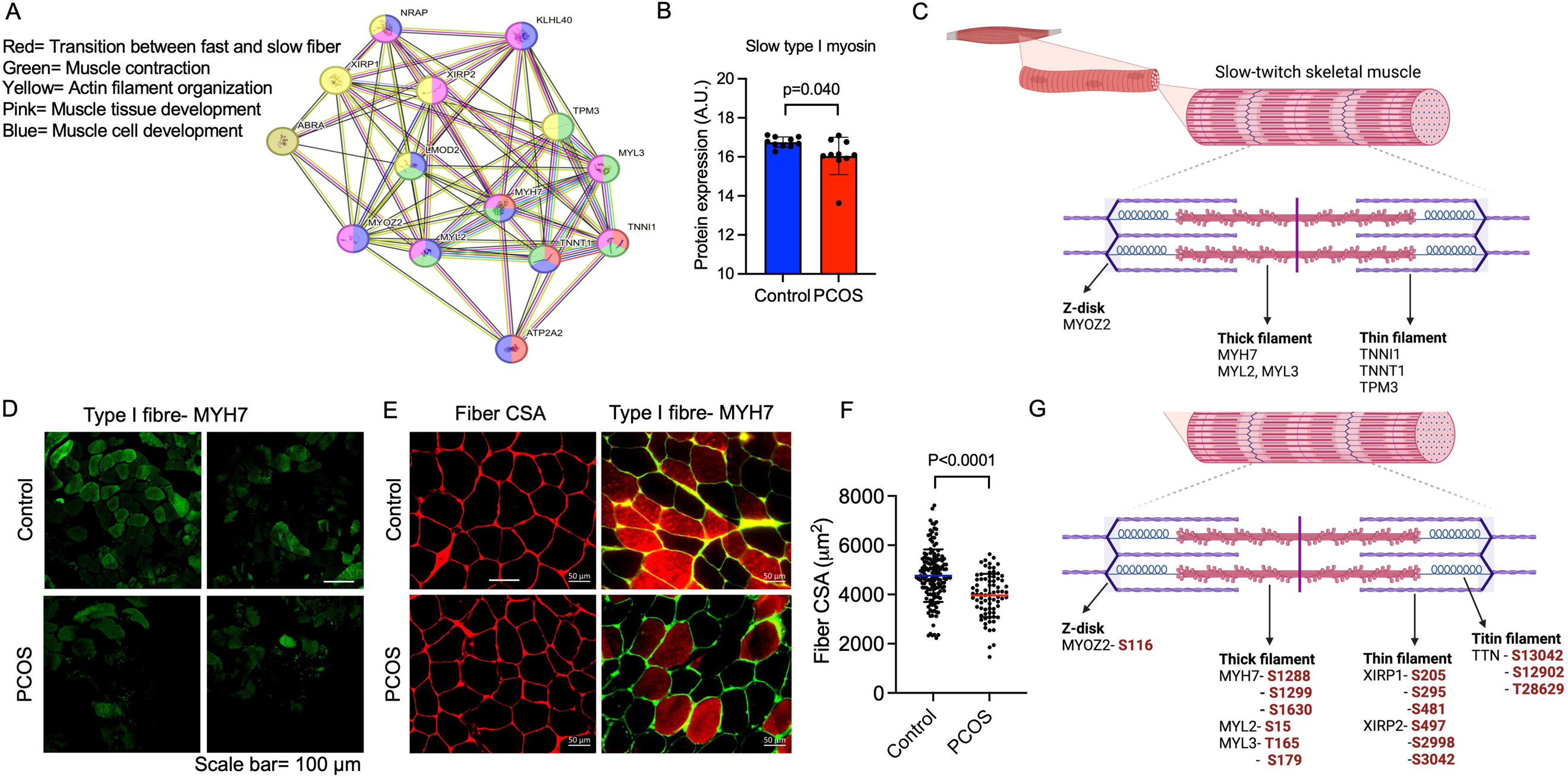
**A)** Protein network on proteins with a lower expression in PCOS skeletal muscle vs. controls. Lines indicate protein-protein associations. **B)** Decreased expression of the slow type I skeletal muscle fibers myosin hevy chain beta (MYH7) in those with PCOS. **C)** Lower expression of proteins in slow-twitch type I muscle fibers in PCOS vs controls (*p*<0.05, log2 fold change <-0.5). **D, E)** Immunoflourescent staining of type I muscle fibers with myosin heavy chain beta, and E) the cell membrane with WGA. **F)** Quantification of fiber cross-sectional area (CSA) in E). G) Differently phosphorylated sites in proteins expressed in muscle filaments (*p*<0.05, log2 fold change ± 0.5).

Then, an androgen-exposed PCOS-like mouse model was used to corroborate that androgen exposure leads to a shift in muscle fiber type. These PCOS-like mice have longer anogenital distance, are in a chronic diestrus phase, and have glucose intolerance (Xiong *et al*., 2022; Ascani *et al*., 2023). These effects are not present in DHT-exposed mice receiving the androgen receptor antagonist flutamide (Ascani *et al*., 2023). DHT-exposed PCOS mice had fewer type I muscle fibers compared to controls (**Fig. 4A-C**). This effect was partly prevented in DHT-exposed mice receiving flutamide, supporting an effect of androgen receptor activation on muscle fiber type (**Fig. 4A, C**). However, although flutamide treatment improves glucose sensitivity in PCOS-like mice, insulin resistance likely also contributes to loss in type I fibers. Moreover, DHT-exposed SkMARKO mice were used to further investigate the contribution of AR-mediated actions in skeletal muscle. While unchallenged SkMARKO mice had fewer type I muscle fibers compared to wt mice (*P*=0.033), they were protected against the androgen-induced type I muscle fiber loss (**Fig. 4B, C**). These data suggest that androgens have direct effects on shifting the muscle fibers towards fewer type I fibers in adult females, which can be prevented by precluding signaling through ARs.

**Fig 4.**
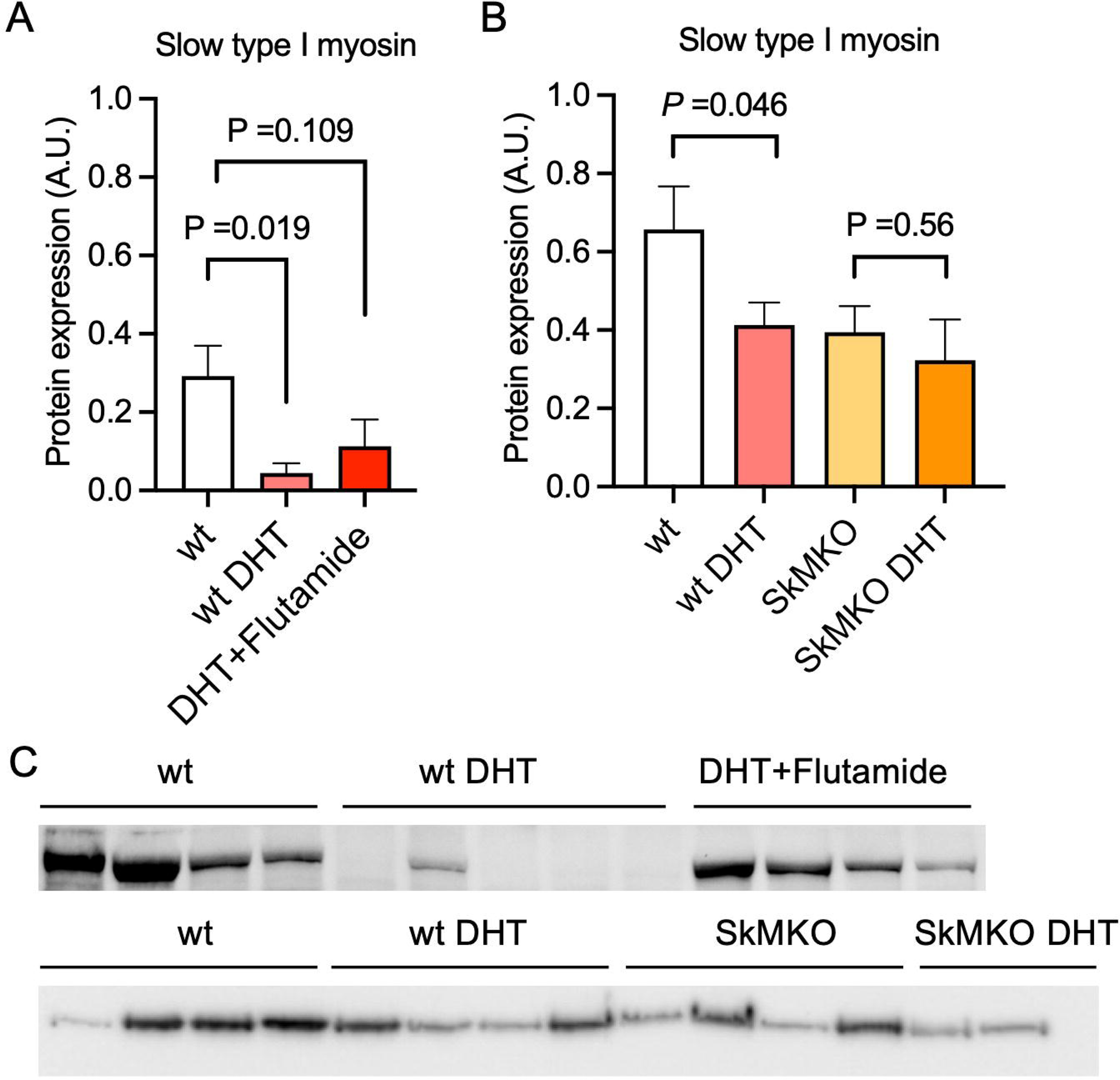
**A)** Decreased expression of the slow type I skeletal muscle fibers myosin heavy chain beta (MYH7) in dihydrotestosterone (DHT)-exposed PCOS-like mice. This effect was partly blocked by the androgen receptor antagonist flutamide (n=5-6/group). **B)** Decreased expression of slow type I skeletal muscle fibers (MYH7) in skeletal muscle-specific androgen receptor knockout mice (SkMARKO) compared to wild type (wt) (p=0.033). DHT exposure did not alter the number of type I fibers in SkMARKO (n=6-8/group) **C)** Representative expression of myosin heavy chain beta. Differences in A) are based on one-way ANOVA and B) on two-way ANOVA, and presented as mean ± SEM. The full raw unedited uncropped blots with the relevant bands clearly labelled are provided as Figure 4-source data 1.

We searched for overlap between the differentially expressed proteins in skeletal muscle in this study and the differentially expressed genes in our previous meta-analysis of gene expression array data (Manti *et al*., 2020). As suspected, the overlap between gene expression and protein levels was small. We found that one upregulated and 12 downregulated genes in muscle biopsies from women with PCOS were also differently expressed at the protein level in this study (**Supplementary file 1c**). Several proteins involved in skeletal muscle contraction were consistently downregulated at the mRNA expression level in muscle tissue from women with PCOS including *MYL3, MYOZ2, TNNT1*, *LMOD2, NRAP,* and *XIRP1* (**Supplementary file 1c**).

We identified 5512 phosphosites in muscle, and 61 sites in 40 unique proteins were differentially phosphorylated in PCOS versus controls (**Supplementary file 1d**), suggesting different protein activity. Eleven of the differently expressed proteins had one or more differently phosphorylated sites, including increased phosphorylation of Ser 130, 382, and 497 in perilipin-1. Many of the proteins in the thick and thin filaments had one or more altered phosphorylation sites (**Fig 3G**). There were no significantly enriched pathways among the differently expressed phosphosites.

### Total protein expression and phosphorylation in adipose tissue

In total, we identified 5000 proteins in adipose tissue but the difference between groups was modest. 21 unique proteins were found to be differentially expressed in adipose tissue from women with PCOS versus controls (*p* < 0.05, and log_2_ FC ≥ 0.5 or ≤ −0.5, **Fig. 5A**). Six proteins were upregulated and 15 were downregulated in women with PCOS and the expression log2 fold-change in expression ranged from 2.1 to −1.6 (**Fig. 5B, Supplementary file 1e**). Several of the upregulated proteins play a role in immune system processes including, immunoglobulins, HLA class I histocompatibility antigen, and sequestosome 1. Sequestosome 1 may also regulate the mitochondrial organization (Poon *et al*., 2021). Three mitochondrial matrix proteins; tRNA pseudouridine synthase A, Enoyl-CoA hydratase domain-containing protein 2, and NAD kinase 2 had an altered expression (**Fig. 5B**), possibly indicating mitochondrial dysfunction. There were three significantly enriched signaling pathways in adipose tissue based on a lower expression of leiomodin-1 and adseverin; actin nucleation (GO:0045010), positive regulation of cytoskeleton organization (GO:0051495), and positive regulation of supramolecular fiber organization (GO:1902905).

**Fig. 5.**
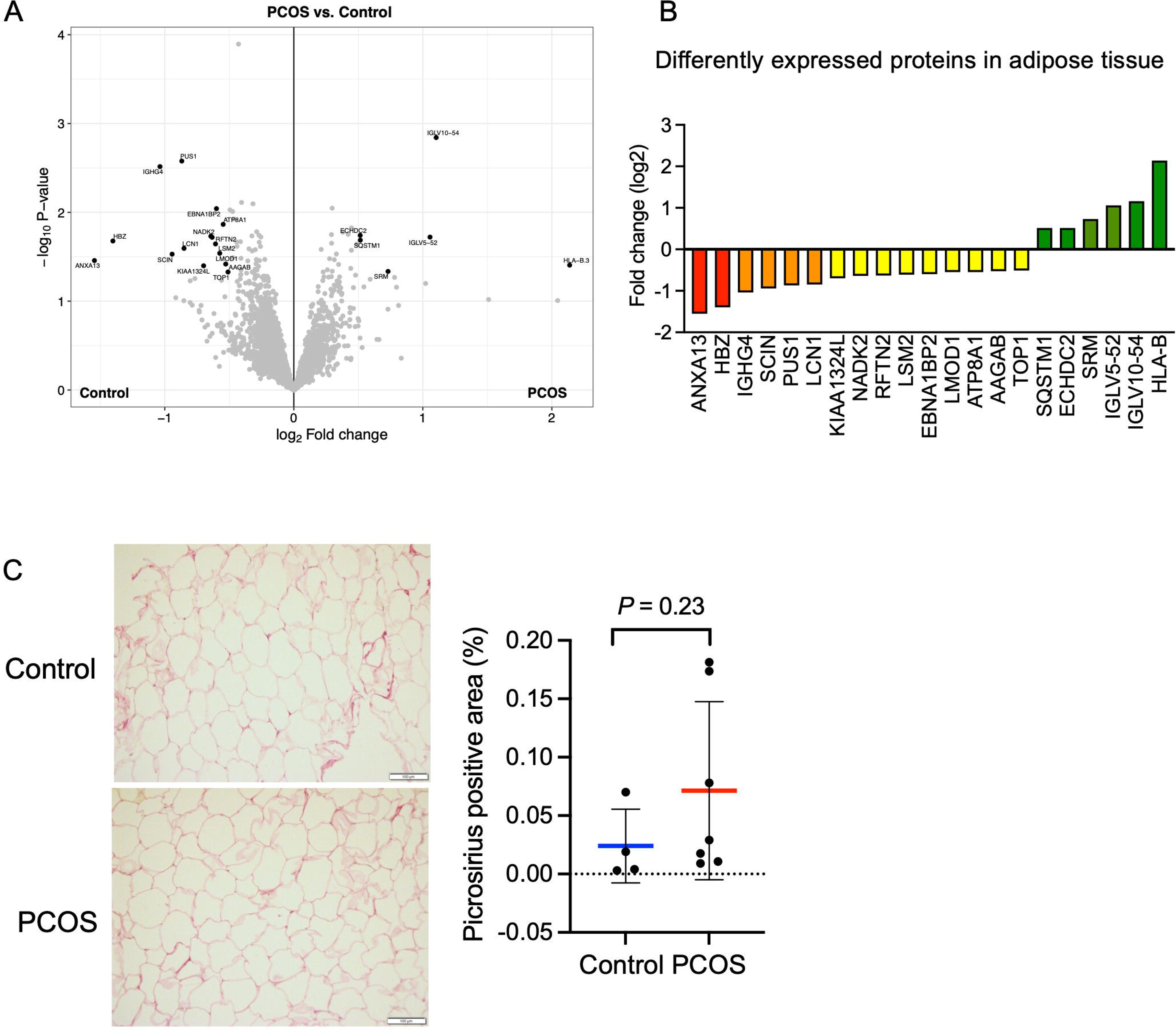
**A)** Volcano plot showing the mean protein log2 fold change in adipose tissue (PCOS vs controls) plotted against the −log10 p value highlighting significantly regulated proteins (black; p<0.05, log2 fold change ± 0.5). n=10/group. **B)** All differentially expressed proteins in adipose tissue from women with PCOS. **C)** Picrosirius red staining of s.c. adipose tissue. Difference between women with PCOS and controls was based on Mann-Whitney U test and is presented as mean ± SD.

Both low-grade inflammation and transforming growth factor beta (TGFβ)-induced fibrosis have been suggested to play a role in the pathophysiology of PCOS (Mancini *et al*., 2021; McIlvenna *et al*., 2021). Dysregulated signaling of TGFβ has been linked to the development of ovarian fibrosis and reproductive dysfunction (Stepto *et al*., 2019; McIlvenna *et al*., 2021). Women with PCOS have elevated levels of circulating TGFβ1 (Raja-Khan *et al*., 2010), which is thought to trigger increased fibrotic mechanisms in other peripheral tissues. However, we did not detect an increased abundance of fibrous collagens in adipose tissue and the fibrillar collagen levels as judged by Picrosirius red staining were similar between groups although the variability was higher in the PCOS group (**Fig. 5C**).

We identified 5734 phosphosites in adipose tissue, of which 39 were differently phosphorylated sites in 34 unique proteins. Ten of these sites had lower phosphorylation (**Supplementary file 1f**). There was no overlap between differently phosphorylated proteins and differently expressed proteins. Perilipin-1 had two phosphorylation sites with higher phosphorylation; Ser 497 and 516 in PCOS adipose tissue compared to controls (**Supplementary file 1f**). Ser 497 showed increased phosphorylation in perilipin-1 in both muscle and adipose tissue. Under adrenergic stimulation, perilipin-1 is phosphorylated at Ser 497 by protein kinase A, which in turn triggers lipolysis by hormone sensitive lipase (HSL) in adipocytes (Marcinkiewicz *et al*., 2006).

### Skeletal muscle protein expression and phosphorylation changes in skeletal muscle after treatment with electrical stimulation

Since long-term electrically stimulated muscle contractions improve glucose regulation and lower androgen levels in women with PCOS (Stener-Victorin *et al*., 2016), we analyzed genome-wide mRNA expression from women with PCOS after treatment to identify changes in skeletal muscle gene expression in response to stimulation. None of the transcripts exhibited changes in expression after FDR correction after 5 weeks of treatment (*q* < 0.05), but 12 transcripts had a FC >1.2 (p<0.05, **Supplementary file 1g**), of which 5 were different collagens. We also analyzed whether the response to electrical stimulation was associated with DNA methylation changes in skeletal muscle. We found that 41,186 (13.8%) of 298,332 analyzed CpG sites had differential methylation in skeletal muscle after treatment (*p* < 0.05), which is almost three times more than expected by chance (*p* < 0.0001, Chi2 test). Of these, 43 CpG sites remained significant after FDR correction (*q* < 0.05, **Supplementary file 1g**). The majority of the sites (74%) showed decreased methylation in response to treatment. The absolute change in DNA methylation in response to treatment ranged from 3 to 14 percentage points.

Since mRNA expression was not significantly regulated in response to repeated electrical stimulations, we investigated whether the effects were regulated at the protein level. We found that 376 unique proteins were changed in skeletal muscle after treatment with electrical stimulation (*p* < 0.05, **Fig. 6A, Supplementary file 1h**). Most proteins were upregulated in women with PCOS after treatment (98%), and the expression log2 fold-change in expression ranged from −1.37 to 2.15. The upregulated proteins were enriched in signaling pathways involved in extracellular matrix (ECM) organization, regulation of TGFβ production, neutrophil-mediated immunity, wound healing, and blood coagulation (**Fig. 6B, Supplementary file 1i**). Proteins involved in ECM organization include 8 different collagens, integrins and TGF-β1 (**Supplementary file 1i**). Collagen 1A1, 1A2 and VCAN were increased after treatment at both gene and protein levels, implying that acupuncture needling elicits a wound healing response. There was a trend towards increased staining of fibrous collagen after treatment, but this was not significant, potentially due to the low number of analyzed good quality sections (**Fig. 6C**). Other upregulated signaling pathways include exocytosis and vesicle transport along the actin filament, muscle contraction and actin filament organization and negative regulation of adenylate cyclase-activating adrenergic signaling. Several effects previously shown to be upregulated after one bout of electroacupuncture were regulated in the opposite direction as pathways involved in negative regulation of angiogenesis, negative regulation of blood vessel morphogenesis, negative regulation of nitric oxide metabolic processes, and regulation of vasoconstriction were enriched. None of the 58 proteins that had a different expression in the PCOS group at baseline were reversed after 5 weeks of treatment.

**Fig. 6.**
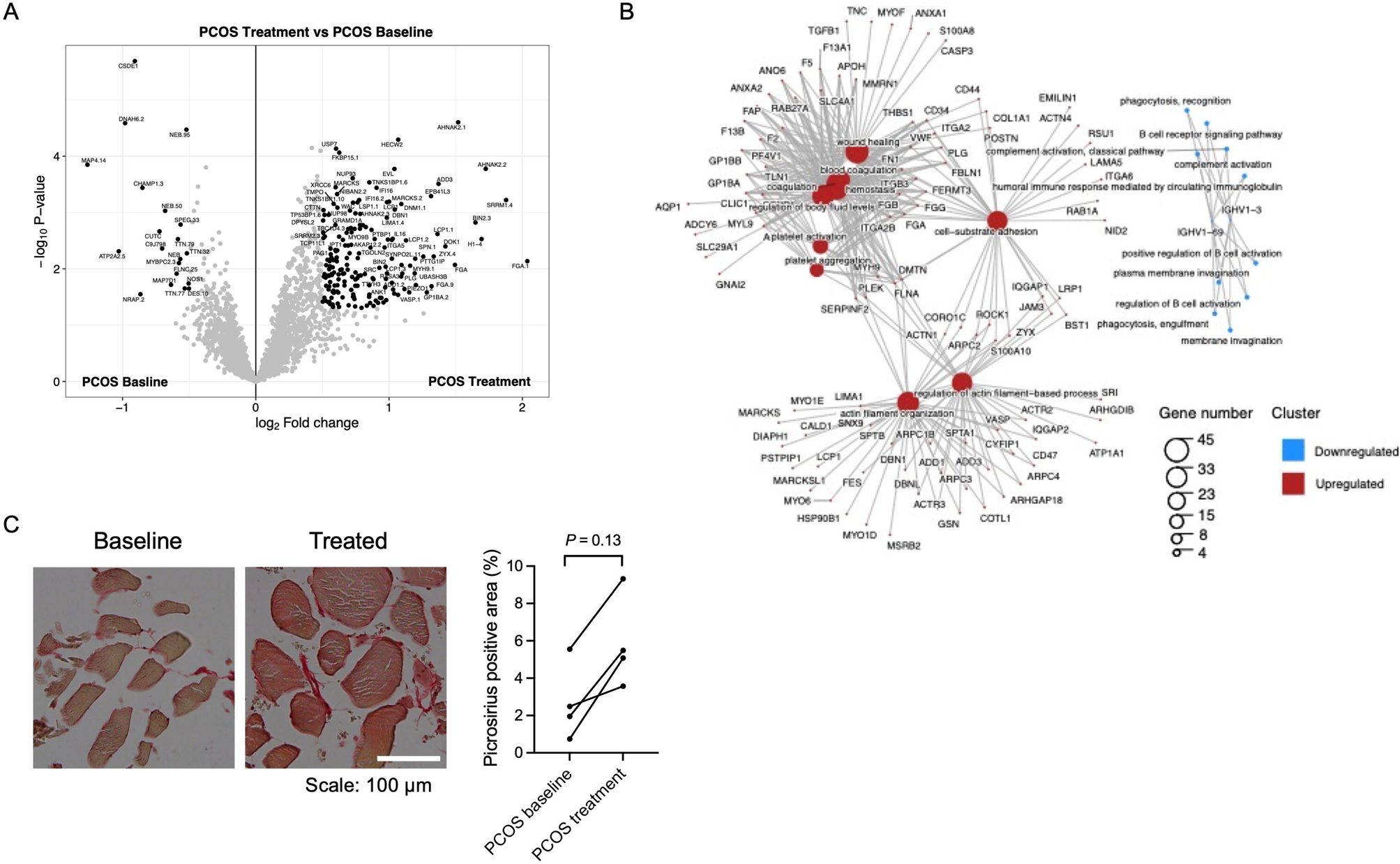
**A)** Volcano plot showing the mean protein log2 fold change in skeletal muscle (treatment vs baseline in PCOS) plotted against the −log10 p value highlighting significantly regulated proteins (black; p<0.05, log2 fold change ± 0.5). n=10/group. **B)** GO terms for biological function of the changed proteins. **C)** Representative pictures and quantification of picrosirius red staining of skeletal muscle before and after treatment with electrical stimulation in the same individual. Changes between baseline and after treatment was based on Wilcoxon signed-rank test.

198 phosphosites in 152 unique proteins showed a changed phosphorylation in response to electrical stimulation. 178 sites had higher phosphorylation, and 46 sites were less phosphorylated (**Supplementary file 1j**). There were no significant enriched pathways among the proteins with differently expressed phosphorylation sites. 38 of the differently expressed proteins had one or more differently phosphorylated sites. These proteins, with changes in both total protein and phosphorylation levels, were enriched in actin filament organization (GO:0007015).

### Total protein expression and phosphorylation in adipose tissue after treatment

Similar to skeletal muscle, long-term electrical stimulations had minimal effects on gene expression in adipose tissue. None of the transcripts exhibited changes in expression after FDR correction (*q* < 0.05) after 5 weeks of treatment, or had a FC >1.2 (data not shown). We found that 23,517 (7.9%) of 298,289 analyzed CpG sites had differential methylation in adipose tissue after 5 weeks of treatment (*p* < 0.05), which is more than expected by chance (*p* < 0.0001, Chi2 test). The majority (63.5%) of these sites showed reduced methylation in response to treatment. One CpG site remained significant after FDR correction (*q* < 0.05, −2.2% points reduced methylation of cg13383058 in the transcription start site of *CD248)*. Therefore, we investigated whether the long-term effects were regulated at the protein level. 61 unique proteins were found to be changed in adipose tissue after electrical stimulation treatment (**Fig. 7A, Supplementary file 1k**). Most of the proteins were upregulated (85%) and 9 were downregulated in women with PCOS after treatment, and the expression log2 fold-change in expression ranged from −0.89 to 1.41. The upregulated enriched signaling pathways include ECM organization and Fc-gamma receptor signaling (**Supplementary file 1l**). In accordance with these findings, 5 weeks of treatment increased the fibrillar collagen content in adipose tissue (**Fig. 7B**). The expression of DNA topoisomerase 1 and leimodulin-1 was lower in women with PCOS than in controls but increased after treatment (**Supplementary file 1k**). Leimodulin-1 is required for proper contractility of smooth muscle cells by mediating nucleation of actin filaments, and myosin regulatory light polypeptide 9 plays an important role in regulating smooth muscle contractile activity. The enzyme prostacyclin synthase, a potent mediator of vasodilation, was also increased and could act on the smooth muscles in the vessel wall. 49 phosphosites in 46 unique proteins showed altered phosphorylation in response to electrical stimulation. All except 4 sites had higher phosphorylation (**Supplementary file 1m**). There were no significantly enriched pathways among the proteins with differently expressed phosphosites.

**Fig. 7.**
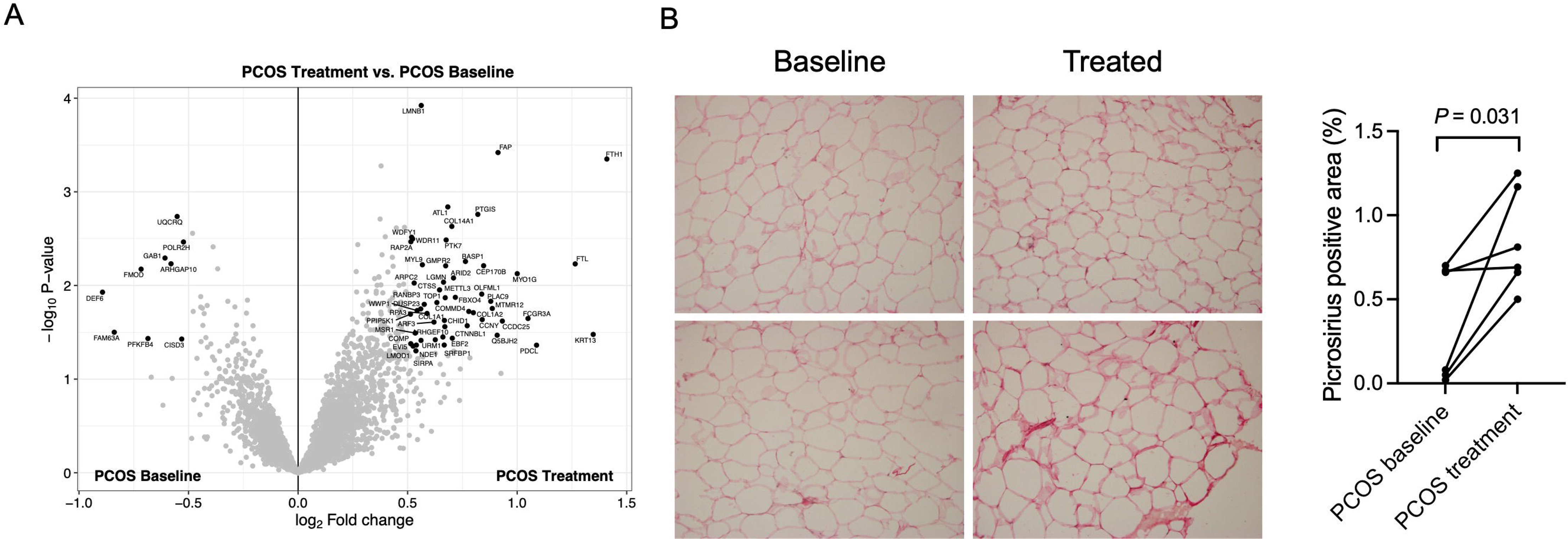
**A)** Volcano plot showing the mean protein log2 fold change in adipose tissue (treatment vs baseline in PCOS) plotted against the −log10 p value highlighting significantly regulated proteins (black; p<0.05, log2 fold change ± 0.5). n=10/group. **B)** Representative pictures and quantification of picrosirius red staining of adipose tissue before and after treatment with electrical stimulation. Changes between baseline and after treatment was based on Wilcoxon signed-rank test.

11 proteins showed higher expression in both skeletal muscle and adipose tissue after treatment; FCGR3A, FAP, PTGIS, COL1A2, COL14A1, COL1A1, MYL9, LMNB1, SIRPA, ARPC2, and RAP2A. These proteins are enriched in ECM organization (GO:0030198), negative regulation of nitric oxide biosynthetic/metabolic processes (GO:0045019, GO:1904406), and Fc-gamma receptor signaling pathways (GO:0038096, GO:0038094, GO:0002431).

## Discussion

### Proteome signature in PCOS

We have profiled the proteome of skeletal muscle and adipose tissue to advance our understanding of the pathophysiology of PCOS. The changes in protein expression in adipose tissue were small, whereas in skeletal muscle of women with PCOS there was a clear downregulation of proteins involved in muscle contraction. The skeletal muscle contains a mixture of slow-twitch oxidative and fast-twitch glycolytic myofibers, which exhibit different physiological properties. Type I, or red fibers, are slow-twitch fatigue resistant muscle fibers that have higher mitochondrial and myoglobin content, and are thus more aerobic than type II fast-twitch fibers. Several proteins specific to or known to be highly expressed in type I muscle fibers were consistently downregulated in women with PCOS. These proteins are located in both the thick and the thin filaments of slow-twitch fibers. These data suggest that there are fewer type I fibers, decreased number and/or smaller, in the PCOS muscle. Unfortunately, we were unable to quantify the number of type I fibers in the entire cohort because of the poor quality of the PCOS muscle biopsies and the unavailability of muscle tissue. We also identified several differently phosphorylated sites in proteins located in the thick and thin filaments, indicating a differential protein activity, since phosphorylation can change the activity of a protein.

Here we show that the signaling pathway important for the transition between fast and slow fibers was downregulated and that individuals with PCOS had lower expression of myosin heavy chain beta (encoded by *MYH7*), which is specific for slow-twitch oxidative type I fibers (Schiaffino & Reggiani, 2011). A decrease in type I fibers has been shown in three different androgen-excess rodent models, including this study (Holmang *et al*., 1990; Shen *et al*., 2019). This effect on type I fibers was partly protected by the coadministration of the androgen receptor antagonist flutamide. Moreover, mice that lack the AR specifically in skeletal muscle were completely protected against this effect. These findings suggest that exaggerated androgen signaling in skeletal muscle directly affects the muscle fiber type composition. While we found a lower abundance of type I muscle fiber in muscle-specific AR knockout females a recent study shows that depleting AR signaling in skeletal muscle does not lead to necrotic fibers, aberrant histology, or changes in key metabolic functions in females (Ghaibour *et al*., 2023). Thus, androgen signaling is likely to be important for normal muscle development but may play a different and dose-dependent role in adulthood. Impaired insulin sensitivity in hyperandrogenic animals is associated with fewer type I fibers and increased type II fibers in skeletal muscles (Holmang *et al*., 1990; Shen *et al*., 2019), features expected to result in reduced insulin sensitivity in this tissue as a higher proportion of oxidative type I fibers leads to better insulin responsiveness (Stuart *et al*., 2013). In line with a higher mitochondrial content in type I fibers, there was a lower expression of several mitochondrial matrix proteins; enoyl acyl carrier protein reductase, 3-oxoacid CoA-transferase 1, enoyl-CoA delta isomerase 1, hydroxyacid-oxoacid transhydrogenase, and acetyl-CoA synthetase 2 (GO:0005759, *q*=0.004) in women with PCOS. Moreover, a lower expression of the mitochondrial acetyl-CoA synthetase 2 correlated with a higher HOMA-IR (Spearman rho=-0.46, *P*=0.04), suggesting that an impaired mitochondrial function contributes to insulin resistance. Moreover, a lower proportion of type I fibers correlated with the severity of insulin resistance in subjects with the metabolic syndrome (Stuart *et al*., 2013). Androgens appear to decrease highly oxidative and insulin-sensitive type I muscle fibers and increase glycolytic and less insulin-sensitive type II fibers in non-athletes, further promoting the development of insulin resistance. However, whether these changes in muscle morphology precede or follow the development of insulin resistance in women with PCOS is not known. Moreover, the sarcoendoplasmic reticulum calcium ATPase 2 (SERCA2a), which is expressed primarily in slow-twitch skeletal muscle, was less abundant in PCOS muscle. The SERCA2a pump transports calcium ions from the cytosol back to the sarcoplasmic reticulum after muscle contraction to keep the cytosolic Ca^2+^ concentration at a low level. Its function is closely related to muscle health and function (Xu & Van Remmen, 2021). There were three sites with lower phosphorylation in SERCA2a, suggesting that lower SERCA2a expression is not only a reflection of decreased type I fibers but also an altered function. Impaired SERCA2 function may lead to increased cytosolic Ca^2+^ concentration, which in turn can impair force production and mitochondrial function in type I fibers (Xu & Van Remmen, 2021). Although serum androgen levels are positively correlated with athletic performance in female athletes (Bermon *et al*., 2018), skeletal muscle contraction and filament sliding pathways were downregulated in muscle biopsies from hyperandrogenic women with overweight/obesity in this study. Thus, androgens may have differential actions on female skeletal muscle function in moderately physically active subjects and female athletes.

Androgens are mainly produced in the ovaries and adrenal glands but there is also a local production in adipose tissue. In overweight/obese women with PCOS, increased AKR1C3 levels mediated increased testosterone generation from androstenedione in subcutaneous adipose tissue, which enhanced lipid storage (O’Reilly *et al*., 2017). Higher expression of AKR1C1 and AKR1C3 in PCOS skeletal muscle in this study could increase local synthesis of androgens via the backdoor pathway, and increase androgenic signaling in skeletal muscle.

The expression of proteins involved in lipid transport and negative regulation of lipid metabolism was increased in the muscles of women with PCOS. Lipid transport was clustered around apolipoprotein C1, C2 and C3. Interestingly, a recent proteomics study shows that serum apolipoprotein C3 levels are higher in insulin resistant women with PCOS compared to insulin sensitive women (Li *et al*., 2020). Could apolipoproteins aid in the deposition of excess fat in skeletal muscle and contribute to lipotoxicity? Fatty acids in skeletal muscle that do not undergo beta-oxidation in the mitochondria inevitably contribute to lipid synthesis. There was indeed a lower expression of various mitochondrial proteins involved in mitochondrial fatty acid beta-oxidation; enoyl acyl carrier protein reductase; enoyl-CoA delta isomerase 1, and acyl-CoA thioesterase 11 (R-HSA-77289, q=0.0008) in PCOS muscle. The most important single fate for these fatty acids is triacylglycerol esterification, and storage in lipid droplets. Perilipins act largely as a scaffold protein on lipid droplets. Perilipin-1 is localized in the periphery of intramuscular lipids, known as extracellular lipids (Gandolfi *et al*., 2011; Zhao *et al*., 2021). In pig, muscle perilipin-1 proteins are localized around lipid droplets in mature and developing adipocytes in correspondence with extra-myocellular lipids (Gandolfi *et al*., 2011; Zhao *et al*., 2021). Therefore, the high expression of peilipin-1 in muscle from women with PCOS is likely a sign of lipotoxicity. Taken together, intramuscular lipid accumulation and type I muscle fiber decline likely contribute to insulin resistance in PCOS muscle.

PCOS is associated with chronically elevated levels of inflammatory markers in the circulation (Orio *et al*., 2005) and low-grade inflammation has been suggested to play a key role in the pathophysiology (Mancini *et al*., 2021). We and others have previously shown that many of the significantly enriched gene expression pathways in PCOS muscle are involved in immune responses or are related to immune diseases in muscle (Skov *et al*., 2007; Nilsson *et al*., 2018). A distinct pattern was the downregulation of a family of genes named the human leukocyte antigen (HLA) complex and downregulation of gene sets associated with inflammatory responses in the PCOS muscle (Manti *et al*., 2020). In this study, the HLA-B was the most downregulated protein, in line with lower gene expression of HLA-B in skeletal muscle (Nilsson *et al*., 2018). The HLA complex helps the immune system distinguish endogenous proteins from proteins made by foreign invaders by making peptide-presenting proteins that are present on the cell surface. When the immune system recognizes the peptides as foreign, this will trigger self-destruction of the infected cell. Numerous HLA alleles have been identified as genetic risk factors for autoimmune thyroid disease, which is increased nearly threefold in women with PCOS (Zeber-Lubecka & Hennig, 2021). The co-occurrence of PCOS and autoimmune thyroid disease has led to the suggestion that PCOS itself may be an autoimmune disorder. At the same time, five different immunoglobulins were upregulated, which contributed to the enrichment of immune system pathways.

In adipose tissue, the difference between groups was small with relatively few differently expressed proteins but several of the proteins with a higher expression play a role in immune system processes including, immunoglobulins and HLA-B. Sequestosome-1 is an immunometabolic protein that is both involved in the activation of nuclear factor kappa-B and regulates mitochondrial functionality by modulating the expression of genes underlying mitochondrial respiration (Poon *et al*., 2021). Alterations in the immune response and immunometabolic pathways are seen in both muscle and adipose tissue, but whether and how these alterations affect, and potentially contribute to the pathophysiology of PCOS remains to be investigated.

### Electrical stimulation-related proteome changes

Regular physical exercise improves insulin sensitivity and is the first-line approach to manage both reproductive and metabolic disturbances in those with PCOS and overweight or obesity, but adherence is low. Electrical stimulations inducing contraction could therefore be an alternative way to ameliorate symptoms in women with PCOS, alongside exercise. Transcriptomic changes in response to one single bout of electrical stimulation mimic the response to one bout of exercise (Benrick *et al*., 2020). Therefore, we hypothesize that long-term treatment with electrical stimulation or exercise also triggers overlapping signaling pathways. Five weeks of repeated treatment with electrical stimulation did not alter mRNA expression but increased the expression of several hundred proteins in skeletal muscle and about 50 proteins in adipose tissue. The most pronounced changes in both tissues were mechanisms involved in wound healing such as the increase in ECM formation and enriched pathways for nitric oxide metabolic process and Fc-gamma receptor signaling pathways. Moreover, wound healing and blood coagulation pathways were upregulated in skeletal muscle, suggesting that repeated needling induces small damages in the tissues that trigger a wound healing response. However, increased expression of ECM-related structural components such as collagens and integrins are not only involved in wound healing, but are also increased in response to muscle contractions as previously shown in response to eccentric exercise and repeated electrical stimulation with electrodes (Mackey *et al*., 2011; Hyldahl *et al*., 2015). We propose that these changes are induced by contraction but we cannot rule out that the ECM-related changes occurred as a direct result of repeated needle insertion. The delayed synthesis of collagens and subsequent strengthening of the ECM structural matrix in response to both exercise and electrical stimulation without needle insertion (Mackey *et al*., 2011; Hyldahl *et al*., 2015), supports the idea that contractions induce remodeling of the ECM that may provide protective adaptation to repeated contractions.

We hypothesized that electrical stimulations mimic the response to repeated exercise. Therefore, we searched for overlap between changes in the proteome following electrical stimulation treatment in this study and a meta-analysis on the exercise-induced proteome in skeletal muscle (Padrao *et al*., 2016). Seven proteins were changed in both conditions; Fibrinogen beta chain, actin, vimentin, annexin A2, moesin, gelsolin, and hypoxanthine-guanine phosphoribosyl-transferase. Actin, vimentin and moesin make up structural parts of the cytoskeleton and all proteins are enriched in the GO terms immune system process (GO:0002376) and positive regulation of metabolic processes (GO:0009893). Current evidence suggests that there are immunometabolic alterations in skeletal muscle from women with PCOS (Skov *et al*., 2007; Nilsson *et al*., 2018; Manti *et al*., 2020; Stepto *et al*., 2020), and increased expression of the abovementioned proteins could cause contraction-induced changes in the immunometabolic response.

Repeated treatment with electrical stimulation improves glucose homeostasis and lowers Hba1c in women with PCOS (Stener-Victorin *et al*., 2016). A not so well characterized, upregulated pathway involves exocytosis and protein transport, which is of interest with regards to glucose transporter 4 (GLUT4) translocation stimulated by AMP kinase (AMPK). The activity of AMPK in muscle increases substantially during contraction and increases glucose transport. AMPK has been linked to at least two mechanisms for the control of vesicle trafficking, namely the regulation of Rab and the generation of phosphatidylinositol 3,5-bisphosphate in the control of GLUT4 translocation (Sylow *et al*., 2017). The Rab GTPase-activating proteins TBC1D1 and TBC1D4 are thought to mediate the effects of AMPK on GLUT4 translocation and glucose transport. At present, it is unclear which specific Rab proteins might be regulated by AMPK downstream of TBC1D1 and TBC1D4, but Rab-13 appears to act downstream of TBC1D4 (Sylow *et al*., 2017). Six Rab proteins showed higher expression after electrical stimulation, including Rab-13, making these proteins potential candidates for regulating glucose uptake by electrically stimulated contractions. The proteome in adipose tissue and skeletal muscle did not show differential expression of proteins with well-known metabolic effects that can easily explain the improvement in Hba1c. This could be due to the time-point when the biopsies were collected, which was within 48 hours of the last treatment when the acute effects of electrical stimulation are no longer present and the long-term changes may be more subtle.

### Limitations

The interpretation of the protein expression in this study is limited by the relatively small number of women (n =10/group), the inclusion of only Caucasian women, and a mix of PCOS phenotypes since we used the Rotterdam diagnostic criteria. Moreover, we cannot distinguish whether the identified dysregulated pathways are responses to PCOS or causal effectors. Some may argue that another limitation is the fact that we cannot identify the changes in protein expression in specific cell types, i.e. adipocytes and myocytes, as the biopsies consist of many different cell types and structures, e.g. nerves, immune cells, vessels and connective tissue. However, this can also be seen as a strength, as no cell acts independently of the cells surrounding it.

### Conclusions

Our findings suggest that highly oxidative and insulin-sensitive type I muscle fibers are decreased in PCOS which in combination with more extramyocellular lipids, may be key factors for insulin resistance in PCOS muscle. In adipose tissue, the difference between groups was small. A 5-week treatment with electrical stimulation triggered a wound healing response in both adipose tissue and skeletal muscle. In addition, remodeling of the ECM can provide protective adaptation to repeated skeletal muscle contractions.

## Additional information

### Data availability statement

The proteomics data have been deposited to the ProteomeXchange Consortium via the Proteomics Identifications (PRIDE) (RRID:SCR_003411) partner repository with the dataset identifier PXD025358. The protein expression analysis is published at https://github.com/GustawEriksson/PCOS. Individual-level methylation and mRNA expression data are not publicly available due to ethical and legal restrictions related to the Swedish Biobanks in Medical Care Act, the Personal Data Act and European Union’s General Data Protection Regulation and Data Protection Act. All other data generated or analyzed during this study are included in the manuscript and supporting files. Raw data can be found at Dryad https://doi.org/10.5061/dryad.wwpzgmsr7

## Data Availability

The proteomics data have been deposited to the ProteomeXchange Consortium via the Proteomics Identifications (PRIDE) (RRID:SCR_003411) partner repository with the dataset identifier PXD025358. The protein expression analysis is published at
https://github.com/GustawEriksson/PCOS. Individual-level methylation and mRNA expression data are not publicly available due to ethical and legal restrictions related to the Swedish Biobanks in Medical Care Act, the Personal Data Act and European Unions General Data Protection Regulation and Data Protection Act. All other data generated or analyzed during this study are included in the manuscript and supporting files. Raw data can be found at Dryad https://doi.org/10.5061/dryad.wwpzgmsr7

## Acknowledgement

For proteomic analysis we thank Britt-Marie Olsson, Johannes Fuchs and Carina Sihlbom at the Proteomics Core Facility, and for the use of its technical equipment and support we thank the Genomics Core Facility, both at Sahlgrenska Academy, University of Gothenburg, Sweden. We are grateful to the Inga-Britt and Arne Lundberg Research Foundation for the donation of the Orbitrap Fusion Tribrid MS instrument. For analysis of array data we thank Michaela Martis at University of Linköping.

## Supporting information

Additional supporting information can be found online in the Supporting Information section at the end of the HTML view of the article. Supporting information files available:

**Figure 4 - Source data 1**. Contains the raw unedited uncropped blots used to generate the figure.

## References

Abdulkhalikova D, Sustarsic A, Vrtacnik Bokal E, Jancar N, Jensterle M & Burnik Papler T. (2022). The Lifestyle Modifications and Endometrial Proteome Changes of Women With Polycystic Ovary Syndrome and Obesity. Front Endocrinol (Lausanne) 13, 888460.

Ascani A, Torstensson S, Risal S, Lu H, Eriksson G, Li C, Teschl S, Menezes J, Sandor K, Ohlsson C, Svensson CI, Karlsson MCI, Stradner MH, Obermayer-Pietsch B & Stener-Victorin E. (2023). The role of B cells in immune cell activation in polycystic ovary syndrome. Elife 12.

Barber TM & Franks S. (2021). Obesity and polycystic ovary syndrome. Clin Endocrinol (Oxf).

Benrick A, Pillon NJ, Nilsson E, Lindgren E, Krook A, Ling C & Stener-Victorin E. (2020). Electroacupuncture Mimics Exercise-Induced Changes in Skeletal Muscle Gene Expression in Women With Polycystic Ovary Syndrome. The Journal of clinical endocrinology and metabolism 105.

Bermon S, Hirschberg AL, Kowalski J & Eklund E. (2018). Serum androgen levels are positively correlated with athletic performance and competition results in elite female athletes. British journal of sports medicine 52, 1531–1532.

Bibikova M, Barnes B, Tsan C, Ho V, Klotzle B, Le JM, Delano D, Zhang L, Schroth GP, Gunderson KL, Fan JB & Shen R. (2011). High density DNA methylation array with single CpG site resolution. Genomics 98, 288–295.

Bolstad BM, Irizarry RA, Astrand M & Speed TP. (2003). A comparison of normalization methods for high density oligonucleotide array data based on variance and bias. Bioinformatics 19, 185–193.

Brower MA, Hai Y, Jones MR, Guo X, Chen YI, Rotter JI, Krauss RM, Legro RS, Azziz R & Goodarzi MO. (2019). Bidirectional Mendelian randomization to explore the causal relationships between body mass index and polycystic ovary syndrome. Hum Reprod 34, 127–136.

Corbould A, Kim YB, Youngren JF, Pender C, Kahn BB, Lee A & Dunaif A. (2005). Insulin resistance in the skeletal muscle of women with PCOS involves intrinsic and acquired defects in insulin signaling. Am J Physiol Endocrinol Metab 288, E1047–1054.

Corton M, Botella-Carretero JI, Lopez JA, Camafeita E, San Millan JL, Escobar-Morreale HF & Peral B. (2008). Proteomic analysis of human omental adipose tissue in the polycystic ovary syndrome using two-dimensional difference gel electrophoresis and mass spectrometry. Hum Reprod 23, 651–661.

Dapas M, Lin FTJ, Nadkarni GN, Sisk R, Legro RS, Urbanek M, Hayes MG & Dunaif A. (2020). Distinct subtypes of polycystic ovary syndrome with novel genetic associations: An unsupervised, phenotypic clustering analysis. PLoS Med 17, e1003132.

Deutsch EW, Bandeira N, Sharma V, Perez-Riverol Y, Carver JJ, Kundu DJ, Garcia-Seisdedos D, Jarnuczak AF, Hewapathirana S, Pullman BS, Wertz J, Sun Z, Kawano S, Okuda S, Watanabe Y, Hermjakob H, MacLean B, MacCoss MJ, Zhu Y, Ishihama Y & Vizcaino JA. (2020). The ProteomeXchange consortium in 2020: enabling ’big data’ approaches in proteomics. Nucleic Acids Res 48, D1145–D1152.

Dumesic DA, Padmanabhan V, Chazenbalk GD & Abbott DH. (2022). Polycystic ovary syndrome as a plausible evolutionary outcome of metabolic adaptation. Reprod Biol Endocrinol 20, 12.

Gandolfi G, Mazzoni M, Zambonelli P, Lalatta-Costerbosa G, Tronca A, Russo V & Davoli R. (2011). Perilipin 1 and perilipin 2 protein localization and gene expression study in skeletal muscles of European cross-breed pigs with different intramuscular fat contents. Meat Sci 88, 631–637.

Ghaibour K, Schuh M, Souali-Crespo S, Chambon C, Charlot A, Rizk J, Rovito D, Rerra AI, Cai Q, Messaddeq N, Zoll J, Duteil D & Metzger D. (2023). Androgen receptor coordinates muscle metabolic and contractile functions. J Cachexia Sarcopenia Muscle 14, 1707–1720.

Group. REA-SPCW. (2004). The Rotterdam ESHRE/ASRM-sponsored PCOS Consensus Workshop Group 2004 Revised 2003 consensus on diagnostic criteria and long-term health risks related to polycystic ovary syndrome (PCOS). Fertil Steril 81, 19–25.

Guo X, Puttabyatappa M, Domino SE & Padmanabhan V. (2020). Developmental programming: Prenatal testosterone-induced changes in epigenetic modulators and gene expression in metabolic tissues of female sheep. Mol Cell Endocrinol 514, 110913.

Holmang A, Svedberg J, Jennische E & Bjorntorp P. (1990). Effects of testosterone on muscle insulin sensitivity and morphology in female rats. Am J Physiol 259, E555–560.

Hutchison SK, Stepto NK, Harrison CL, Moran LJ, Strauss BJ & Teede HJ. (2011). Effects of exercise on insulin resistance and body composition in overweight and obese women with and without polycystic ovary syndrome. The Journal of clinical endocrinology and metabolism 96, E48–56.

Hyldahl RD, Nelson B, Xin L, Welling T, Groscost L, Hubal MJ, Chipkin S, Clarkson PM & Parcell AC. (2015). Extracellular matrix remodeling and its contribution to protective adaptation following lengthening contractions in human muscle. FASEB journal : official publication of the Federation of American Societies for Experimental Biology 29, 2894–2904.

Insenser M, Montes-Nieto R, Martinez-Garcia MA & Escobar-Morreale HF. (2016). A nontargeted study of muscle proteome in severely obese women with androgen excess compared with severely obese men and nonhyperandrogenic women. Eur J Endocrinol 174, 389–398.

Jedel E, Labrie F, Oden A, Holm G, Nilsson L, Janson PO, Lind AK, Ohlsson C & Stener-Victorin E. (2011). Impact of electro-acupuncture and physical exercise on hyperandrogenism and oligo/amenorrhea in women with polycystic ovary syndrome: a randomized controlled trial. Am J Physiol Endocrinol Metab 300, E37–45.

Joham AE, Norman RJ, Stener-Victorin E, Legro RS, Franks S, Moran LJ, Boyle J & Teede HJ. (2022). Polycystic ovary syndrome. Lancet Diabetes Endocrinol 10, 668–680.

Johansson J, Redman L, Veldhuis PP, Sazonova A, Labrie F, Holm G, Johannsson G & Stener-Victorin E. (2013). Acupuncture for ovulation induction in polycystic ovary syndrome: a randomized controlled trial. Am J Physiol Endocrinol Metab 304, E934–943.

Johnson WE, Li C & Rabinovic A. (2007). Adjusting batch effects in microarray expression data using empirical Bayes methods. Biostatistics 8, 118–127.

Kakoly NS, Earnest A, Teede HJ, Moran LJ & Joham AE. (2019). The Impact of Obesity on the Incidence of Type 2 Diabetes Among Women With Polycystic Ovary Syndrome. Diabetes Care 42, 560–567.

Kokosar M, Benrick A, Perfilyev A, Fornes R, Nilsson E, Maliqueo M, Behre CJ, Sazonova A, Ohlsson C, Ling C & Stener-Victorin E. (2016). Epigenetic and Transcriptional Alterations in Human Adipose Tissue of Polycystic Ovary Syndrome. Sci Rep 6, 22883.

Li L, Zhang J, Zeng J, Liao B, Peng X, Li T, Li J, Tan Q, Li X, Yang Y, Chen Z & Liang Z. (2020). Proteomics analysis of potential serum biomarkers for insulin resistance in patients with polycystic ovary syndrome. Int J Mol Med 45, 1409–1416.

Liu Q, Zhu Z, Kraft P, Deng Q, Stener-Victorin E & Jiang X. (2022). Genomic correlation, shared loci, and causal relationship between obesity and polycystic ovary syndrome: a large-scale genome-wide cross-trait analysis. BMC Med 20, 66.

Mackey AL, Brandstetter S, Schjerling P, Bojsen-Moller J, Qvortrup K, Pedersen MM, Doessing S, Kjaer M, Magnusson SP & Langberg H. (2011). Sequenced response of extracellular matrix deadhesion and fibrotic regulators after muscle damage is involved in protection against future injury in human skeletal muscle. FASEB journal : official publication of the Federation of American Societies for Experimental Biology 25, 1943–1959.

Mancini A, Bruno C, Vergani E, d’Abate C, Giacchi E & Silvestrini A. (2021). Oxidative Stress and Low-Grade Inflammation in Polycystic Ovary Syndrome: Controversies and New Insights. Int J Mol Sci 22.

Manneras-Holm L, Benrick A & Stener-Victorin E. (2014). Gene expression in subcutaneous adipose tissue differs in women with polycystic ovary syndrome and controls matched pair-wise for age, body weight, and body mass index. Adipocyte 3, 190–196.

Manneras-Holm L, Leonhardt H, Kullberg J, Jennische E, Oden A, Holm G, Hellstrom M, Lonn L, Olivecrona G, Stener-Victorin E & Lonn M. (2011). Adipose tissue has aberrant morphology and function in PCOS: enlarged adipocytes and low serum adiponectin, but not circulating sex steroids, are strongly associated with insulin resistance. The Journal of clinical endocrinology and metabolism 96, E304–311.

Manti M, Stener-Victorin E & Benrick A. (2020). Skeletal Muscle Immunometabolism in Women With Polycystic Ovary Syndrome: A Meta-Analysis. Front Physiol 11, 573505.

Marcinkiewicz A, Gauthier D, Garcia A & Brasaemle DL. (2006). The phosphorylation of serine 492 of perilipin a directs lipid droplet fragmentation and dispersion. The Journal of biological chemistry 281, 11901–11909.

McIlvenna LC, Patten RK, McAinch AJ, Rodgers RJ, Stepto NK & Moreno-Asso A. (2021). Transforming Growth Factor Beta 1 Alters Glucose Uptake but Not Insulin Signalling in Human Primary Myotubes From Women With and Without Polycystic Ovary Syndrome. Front Endocrinol (Lausanne) 12, 732338.

Montes-Nieto R, Insenser M, Martinez-Garcia MA & Escobar-Morreale HF. (2013). A nontargeted proteomic study of the influence of androgen excess on human visceral and subcutaneous adipose tissue proteomes. The Journal of clinical endocrinology and metabolism 98, E576–585.

Moreno-Asso A, Altintas A, McIlvenna LC, Patten RK, Botella J, McAinch AJ, Rodgers RJ, Barres R & Stepto NK. (2021). Non-cell autonomous mechanisms control mitochondrial gene dysregulation in polycystic ovary syndrome. Journal of molecular endocrinology 68, 63–76.

Nilsson E, Benrick A, Kokosar M, Krook A, Lindgren E, Kallman T, Martis MM, Hojlund K, Ling C & Stener-Victorin E. (2018). Transcriptional and Epigenetic Changes Influencing Skeletal Muscle Metabolism in Women With Polycystic Ovary Syndrome. The Journal of clinical endocrinology and metabolism 103, 4465–4477.

O’Reilly MW, Kempegowda P, Walsh M, Taylor AE, Manolopoulos KN, Allwood JW, Semple RK, Hebenstreit D, Dunn WB, Tomlinson JW & Arlt W. (2017). AKR1C3-Mediated Adipose Androgen Generation Drives Lipotoxicity in Women With Polycystic Ovary Syndrome. The Journal of clinical endocrinology and metabolism 102, 3327–3339.

Orio F, Jr., Palomba S, Cascella T, Di Biase S, Manguso F, Tauchmanova L, Nardo LG, Labella D, Savastano S, Russo T, Zullo F, Colao A & Lombardi G. (2005). The increase of leukocytes as a new putative marker of low-grade chronic inflammation and early cardiovascular risk in polycystic ovary syndrome. The Journal of clinical endocrinology and metabolism 90, 2–5.

Padrao AI, Ferreira R, Amado F, Vitorino R & Duarte JA. (2016). Uncovering the exercise-related proteome signature in skeletal muscle. Proteomics 16, 816–830.

Poon A, Saini H, Sethi S, O’Sullivan GA, Plun-Favreau H, Wray S, Dawson LA & McCarthy JM. (2021). The role of SQSTM1 (p62) in mitochondrial function and clearance in human cortical neurons. Stem Cell Reports 16, 1276–1289.

Raja-Khan N, Kunselman AR, Demers LM, Ewens KG, Spielman RS & Legro RS. (2010). A variant in the fibrillin-3 gene is associated with TGF-beta and inhibin B levels in women with polycystic ovary syndrome. Fertility and sterility 94, 2916–2919.

Ronn T, Volkov P, Gillberg L, Kokosar M, Perfilyev A, Jacobsen AL, Jorgensen SW, Brons C, Jansson PA, Eriksson KF, Pedersen O, Hansen T, Groop L, Stener-Victorin E, Vaag A, Nilsson E & Ling C. (2015). Impact of age, BMI and HbA1c levels on the genome-wide DNA methylation and mRNA expression patterns in human adipose tissue and identification of epigenetic biomarkers in blood. Human molecular genetics 24, 3792–3813.

Schiaffino S & Reggiani C. (2011). Fiber types in mammalian skeletal muscles. Physiol Rev 91, 1447–1531.

Shen Q, Bi H, Yu F, Fan L, Zhu M, Jia X & Kang J. (2019). Nontargeted metabolomic analysis of skeletal muscle in a dehydroepiandrosterone-induced mouse model of polycystic ovary syndrome. Mol Reprod Dev 86, 370–378.

Skov V, Glintborg D, Knudsen S, Jensen T, Kruse TA, Tan Q, Brusgaard K, Beck-Nielsen H & Hojlund K. (2007). Reduced expression of nuclear-encoded genes involved in mitochondrial oxidative metabolism in skeletal muscle of insulin-resistant women with polycystic ovary syndrome. Diabetes 56, 2349–2355.

Smyth GK. (2004). Linear models and empirical bayes methods for assessing differential expression in microarray experiments. Stat Appl Genet Mol Biol 3, Article3.

Stener-Victorin E, Jedel E, Janson PO & Sverrisdottir YB. (2009). Low-frequency electroacupuncture and physical exercise decrease high muscle sympathetic nerve activity in polycystic ovary syndrome. Am J Physiol Regul Integr Comp Physiol 297, R387–395.

Stener-Victorin E, Maliqueo M, Soligo M, Protto V, Manni L, Jerlhag E, Kokosar M, Sazonova A, Behre CJ, Lind M, Ohlsson C, Hojlund K & Benrick A. (2016). Changes in HbA1c and circulating and adipose tissue androgen levels in overweight-obese women with polycystic ovary syndrome in response to electroacupuncture. Obes Sci Pract 2, 426–435.

Stepto NK, Hiam D, Gibson-Helm M, Cassar S, Harrison CL, Hutchison SK, Joham AE, Canny BJ, Moreno-Asso A, Strauss BJ, Hatzirodos N, Rodgers RJ & Teede HJ. (2020). Exercise and insulin resistance in PCOS: muscle insulin signalling and fibrosis. Endocr Connect 9, 346–359.

Stepto NK, Moreno-Asso A, McIlvenna LC, Walters KA & Rodgers RJ. (2019). Molecular Mechanisms of Insulin Resistance in Polycystic Ovary Syndrome: Unraveling the Conundrum in Skeletal Muscle? The Journal of clinical endocrinology and metabolism 104, 5372–5381.

Stuart CA, McCurry MP, Marino A, South MA, Howell ME, Layne AS, Ramsey MW & Stone MH. (2013). Slow-twitch fiber proportion in skeletal muscle correlates with insulin responsiveness. The Journal of clinical endocrinology and metabolism 98, 2027–2036.

Sylow L, Kleinert M, Richter EA & Jensen TE. (2017). Exercise-stimulated glucose uptake - regulation and implications for glycaemic control. Nat Rev Endocrinol 13, 133–148.

Tiwari N, Pasrija S & Jain S. (2019). Randomised controlled trial to study the efficacy of exercise with and without metformin on women with polycystic ovary syndrome. Eur J Obstet Gynecol Reprod Biol 234, 149–154.

Villa J & Pratley RE. (2011). Adipose tissue dysfunction in polycystic ovary syndrome. Curr Diab Rep 11, 179–184.

Wang W, Jiang Q, Niu Y, Ding Q, Yang X, Zheng Y, Hao J & Wei D. (2022). Proteomics and bioinformatics analysis of follicular fluid from patients with polycystic ovary syndrome. Front Mol Biosci 9, 956406.

Xiong T, Rodriguez Paris V, Edwards MC, Hu Y, Cochran BJ, Rye KA, Ledger WL, Padmanabhan V, Handelsman DJ, Gilchrist RB & Walters KA. (2022). Androgen signaling in adipose tissue, but less likely skeletal muscle, mediates development of metabolic traits in a PCOS mouse model. Am J Physiol Endocrinol Metab 323, E145–E158.

Xu H & Van Remmen H. (2021). The SarcoEndoplasmic Reticulum Calcium ATPase (SERCA) pump: a potential target for intervention in aging and skeletal muscle pathologies. Skelet Muscle 11, 25.

Zeber-Lubecka N & Hennig EE. (2021). Genetic Susceptibility to Joint Occurrence of Polycystic Ovary Syndrome and Hashimoto’s Thyroiditis: How Far Is Our Understanding? Front Immunol 12, 606620.

Zhang X, Smits AH, van Tilburg GB, Ovaa H, Huber W & Vermeulen M. (2018). Proteome-wide identification of ubiquitin interactions using UbIA-MS. Nat Protoc 13, 530–550.

Zhao Y, Albrecht E, Li Z, Schregel J, Sciascia QL, Metges CC & Maak S. (2021). Distinct Roles of Perilipins in the Intramuscular Deposition of Lipids in Glutamine-Supplemented, Low-, and Normal-Birth-Weight Piglets. Front Vet Sci 8, 633898.

Zhao Y, Xu Y, Wang X, Xu L, Chen J, Gao C, Wu C, Pan D, Zhang Q, Zhou J, Chen R, Wang Z, Zhao H, You L, Cao Y, Li Z & Shi Y. (2020). Body Mass Index and Polycystic Ovary Syndrome: A 2-Sample Bidirectional Mendelian Randomization Study. The Journal of clinical endocrinology and metabolism 105.

